# Concurrent and predictive validity of dynamic assessments of word reading in young children: A systematic review and meta-analysis

**DOI:** 10.1101/2022.09.19.22279942

**Authors:** Emily Wood, Kereisha Biggs, Monika Molnar

**Affiliations:** Department of Speech-Language Pathology, University of Toronto; Rehabilitation Sciences Institute, University of Toronto

**Keywords:** Dynamic assessment, Static assessment, Concurrent validity, Predictive Validity, Early literacy, Reading, Decoding, Phonological awareness, Alphabetic principle, Bilingual, At-risk

## Abstract

Early evaluation of word reading skills is an important step in understanding and predicting children’s future literacy abilities. Traditionally, word reading evaluations are conducted using ‘static’ assessments (SA), which measure a child’s acquired knowledge and are prone to floor effects. Additionally, many of these tools are developed exclusively for English monolinguals, and therefore cannot be used equitably to evaluate the abilities of bilingual children. Dynamic assessment (DA), which evaluates the ability to learn a skill, is a potentially more equitable alternative. To establish that use of DAs is a valid alternative to traditional SAs, their concurrent agreement with gold standard SA measures and their predictive agreement with later word reading outcomes should be considered. In line with this, the primary objective of this systematic review and meta-analysis is to examine the concurrent and predictive validity of DAs of word reading skills. Two secondary objectives are (i) to address which types of word reading DAs (phonological awareness, sound-symbol knowledge, or decoding) demonstrate the strongest relationships with equivalent concurrent static measures and later word reading outcomes, and (ii) to consider for which populations, defined by language status (monolingual vs. bilingual vs. mixed) and reading status (typically developing vs. at-risk vs. mixed) these DAs are valid. Thirty-four studies from 32 papers were identified through searching 5 databases, and the grey literature. Included studies provided a correlation between a DA and concurrent SA, or a DA and a later word reading outcome measure. Regarding concurrent validity, we observed a strong relationship between DAs and SAs in general (r=.60); however, subgroup analyses indicate that DAs of decoding (r=.54) and phonological awareness (r=.73) measures demonstrate greater strength of correlation with their static counterparts, compared to DAs of sound-symbol knowledge (r=.34). In terms of predictive validity, we observed a similarly strong relationship between DAs and word reading outcome measures (r=.57), independently of the type of measure. Subgroup analyses conducted based on participant language status suggested that there are significant differences between mean effect sizes for monolingual, bilingual and mixed language groups in terms of DAs’ concurrent validity with SAs, but no significant differences for predictive validity with word reading outcome measures. There were also no significant differences between mean effect sizes for at-risk, typically developing, or mixed groups in terms of DAs concurrent validity with SAs or predictive validity with word reading outcome measures. Results provide preliminary evidence to suggest that DAs of phonological awareness and decoding skills are a valid alternative to SAs of equivalent constructs and are valid for the future prediction of word reading outcomes across population groups regardless of their language or reading status.

## Introduction

### Literacy

Literacy is the ability to understand, interpret, create, and communicate with information in print, is necessary for social, academic, professional, and personal success and is considered a fundamental human right (Montoya, 2018; Moretti & Frandell, 2013). In 2017, the United Nations Educational, Scientific and Cultural Organization (UNESCO) reported that more than 50% of children worldwide had literacy difficulties, and problem that has been exacerbated by the COVID-19 pandemic (UNESCO, 2021; Aurini & Davies, 2021). Inadequate literacy skills are associated with negative life outcomes, such as poor physical (Wolf et al., 2005) and mental health (Daniel et al., 2006), reduced academic attainment (Ritchie & Bates, 2013), restricted socioeconomic mobility (Barwick & Siegel, 1996), and increased rates of poverty, homelessness, and incarceration (Shelley-Tremblay et al., 2007).

Research has established that early identification is key in mitigating future literacy difficulties and their associated adverse effects (Lundberg, 1994; Ontario Human Rights Commission, 2022). The construct of word reading skills in this review was informed by the subskills that comprise word recognition ability in the evidence-based reading model - Scarborough’s Reading Rope (2001). This model identifies three subskills (i) phonological awareness (PA)– the ability to identify and manipulate parts of speech, (ii) knowledge of the alphabetic principle, or sound-symbol knowledge (SSK)-the ability to recognize the systematic relationship between symbol(s) (letter) and the sound(s) they represent in print and (iii) sight recognition – the ability to apply PA and SSK skills to rapidly and automatically read or decode words, as essential for success in early word recognition and word reading (Scarborough, 2001). While these early word reading skills are foundational, they alone are not sufficient for developing reading ability, as children must also comprehend what they decode. However, word recognition skills contribute more greatly to initial reading success than comprehension skills (Scarborough, 1998). Numerous studies demonstrated that word recognition skills play a significant role in early reading development and prediction of future reading outcomes (e.g., Catts et al., 2005; Hogan et al., 2005). Given that this review is focused on assessments used to evaluate children who are still in the stages of learning to read and decode (i.e., ages 4-9) our goal is to examine the validity of DAs of word reading ability – specifically, phonological awareness, sound-symbol knowledge, and decoding. Examining early reading comprehension is beyond the scope of the current review.

### Traditional Static Word Reading Assessment

A range of traditional standardized assessment tools (e.g., The Comprehensive Test of Phonological Processing-2 (CTOPP-2), Wagner et al., 2013; Phonological Awareness Test-2: Normative Update (PAT-2:NU), Robertson & Salter, 2017; Test of Auditory Processing Skills-4 (TAPS-4), Martin et al., 2018) are commonly used by clinicians, educators, and researchers, to assess early word reading skills. These so-called “static assessments” (SAs) quantify a child’s acquired knowledge and either compare their performance to same-aged peers through norm-referenced scores or determine if they can demonstrate an expected skill by evaluating whether they can complete a specific criterion-referenced task (Grigorenko & Sternberg, 1998). Two potential difficulties are associated with these traditional SAs. First, many of these assessments have been developed exclusively for use with English monolinguals. This English, monolingual-centric focus in test development is at odds with the global population as about half of individuals speak at least two languages (Grosjean, 2010). From both a research and clinical perspective, it is inequitable to use English monolingual assessments to evaluate the skills of a bilingual child. Not only might these tools be linguistically and culturally inappropriate, but testers also cannot be sure if performance differences are due to lack of ability, or language effects (bilingual vs. monolingual) that mask a child’s capacity to perform the skill. A bilingual kindergarten student who speaks Tamil at home and begins learning English in school is likely to perform much more poorly than a child who has grown up speaking exclusively English on a static English test of letter-sound knowledge not because they lack the ability to learn letter sound relationships, but either because of limited exposure to English letters and sounds relative to English monolingual peers; or because of an inability to understand English instructions or a lack of familiarity with Eurocentric or Western assessment practices and ways of measuring knowledge. These cultural and language effects associated with SAs often result in misidentification of reading difficulties in bilingual populations (Bedore & Peña, 2008; Petersen & Gillam, 2015).

The second potential issue associated with SAs is floor effects (Catts et al., 2009). Floor effects are commonly observed in traditional assessment of word reading because these tools attempt to quantify a child’s acquired ability in an area with which they have limited to no experience. Many kindergarten-aged children, even English monolinguals for whom the tests are developed, perform poorly on SAs of word reading ability at the start of the school year, simply because they have had limited experience with these types of tasks. When most children who take a test perform poorly, examiners are unable to differentiate those who truly are at risk for future reading challenges from those who are so-called false positives; students with limited previous experience who will quickly catch up, or those whose linguistic and cultural experiences did not permit them to demonstrate their capabilities on the test. Traditional tests may underestimate the capabilities of a child with minimal literacy experiences, by suggesting that their current lack of knowledge is predictive of their future ability.

### Dynamic Assessment as an Alternative

Dynamic assessment (DA) is an alternative to the traditional SA paradigm. Unlike SAs which evaluate acquired knowledge DAs attempt to measure the ability to learn. This is achieved through interactive testing which can incorporate teaching, scaffolding, prompting and feedback, resulting in an assessment that more closely resembles real world learning experiences (Grigorenko & Sternberg, 1998; Poehner, 2008). Interest in DAs of word reading abilities has been steadily growing in the research community, (e.g., Cho et al., 2017; Gellert & Elbro, 2017a; Petersen et al., 2016), but their clinical use remains limited, with professionals like Speech-Language Pathologists (SLPs) continuing to favour use of SAs (e.g., Arias & Friberg, 2017; D’Souza et al., 2012).

A potential reason for this lack of uptake in use of DA could be that these tools, which are typically less structured than SAs and often unstandardized, are not viewed as a psychometrically valid alternative to SAs. Examining DAs concurrent validity with traditional SAs and predictive validity with word reading outcome measures will contribute to understanding DAs potential as an alternative method for evaluating and predicting literacy abilities. This is the overarching goal of the current systematic review and meta-analyses.

### Previous Reviews of Dynamic Assessment

Several prior reviews have evaluated the use and validity of DAs. Here, we focus on three that address DAs of literacy. Caffrey et al.’s 2008 systematic review and correlational meta-analysis offered support for the predictive validity of DAs, but their analyses did not differentiate between the various included domains (e.g., math, cognition and reading). Furthermore, at-risk and bilingual children were merged in their analyses, despite bilingualism not being an inherent risk factor for reading disorders or any disorders in general. In two subsequent systematic review papers, Dixon et al. (2022a, 2022b) investigated whether DAs can uniquely predict variance in the growth of a child’s reading development beyond SAs, and whether DAs can act as a viable alternative to diagnosing reading disorders in children. In these reviews, the assessment of specific skills that typically predict future reading success were considered, such as phonological awareness, decoding, and morphological awareness. Their findings suggest that DAs of phonological awareness, decoding, and morphological awareness can account for variance in the growth of different outcome measures, ranging from 1-33% (Dixon et al., 2022a); and that DAs can account for unique variance in predicting later reading disorder, particularly when the test construct is similar to reading (e.g., a DA of decoding better predicts word reading outcomes than a DA of working memory) and when predicting abilities proximally vs distally (e.g., in early vs later school years; Dixon et al., 2022b).

While findings from these three previous reviews are promising, there are gaps that remain unaddressed. First, no prior review has examined the concurrent validity of DAs of word reading skills, qualitatively or quantitatively. Evaluating a tool’s concurrent agreement with gold standard measures is a key component of establishing its criterion related validity and determining whether it can serve as a legitimate alternative to established assessments. Furthermore, while the Caffrey et al., (2008) and Dixon et al., (2022a, 2022b) studies considered predictive validity of DAs, neither conducted quantitative analyses which systematically examined (i) the predictive validity of distinct types of word reading word reading DAs or (ii) the validity of word reading DAs for use with populations based on their language (monolingual vs. bilingual) and reading status (at-risk vs. typically developing). All of which are important for understanding the validity of DAs across various populations and contexts.

### The current study

This systematic review and correlational meta-analysis will address these gaps. First, we will quantitatively evaluate the concurrent validity of DAs of word reading skills (phonological awareness, sound-symbol knowledge, and decoding), with their equivalent SAs, as well as the predictive validity of these same word reading DAs with later word reading outcomes, defined in this review as single word or nonword reading. Secondly, we will investigate whether these DAs of word reading skills demonstrate consistent concurrent and predictive validity across population groups, defined by language status (monolingual vs. bilingual), and reading status (at-risk vs. typically developing). Finally, unlike all previous reviews of DA, we will conduct an extensive grey literature search, to reduce potential publication bias, and will include studies published in languages other than English (Spanish, French).

## Method

Methods and analyses were planned a priori and outlined in a systematic review and meta-analysis protocol. This protocol was preregistered on Open Science Framework and is available at online at https://osf.io/bcghx/ (Wood & Molnar, 2022).

### Research Questions

This systematic review and meta-analyses was designed to address the following questions:

**1.A)** Do dynamic assessments of word reading skills (phonological awareness, sound-symbol knowledge, and decoding), demonstrate ***concurrent validity*** with static assessments of word reading skills (PA, SSK, decoding) across all populations?

**1.B)** Do dynamic assessments of word reading skills demonstrate ***predictive validity*** with reading outcome measures (single word reading) across all populations?

**2. A)** Do dynamic assessments of word reading skills demonstrate concurrent validity with static assessments of word reading skills and predictive validity with word and nonword reading outcomes within population groups defined by their ***language status*** (monolingual vs. Bilingual)?

**2. B)** Do dynamic assessments of word reading skills demonstrate concurrent validity with static assessments of word reading skills and predictive validity with word reading outcome measures within population groups defined by their ***reading status*** (at-risk vs. typically developing)?

### Eligibility Criteria

Study inclusion criteria were decided upon in advance and outlined in the systematic review and meta-analysis screening protocol (Wood & Molnar, 2022):

i. Only primary research articles were included. Case reports, commentaries and editorials were excluded, as well as systematic reviews and books or book chapters. Articles published in peer-reviewed journals, and unpublished grey literature from preprint repositories, and reports or dissertations were included.
ii. Given that the focus of the current paper is early word reading skills, only studies that evaluated children with a mean age of 4;0 – 10;0, whose participants were either typically-developing, at-risk for reading difficulty, diagnosed with a reading disorder and monolingual or bi/multilingual were included. Studies conducted with adults or with children with developmental difficulties (e.g., developmental language disorder, autism spectrum disorder, hearing difficulty) were excluded.
iii. All included studies used a DA of word reading skills and (i) a SA of word reading skills at the same time point, AND/OR (ii) a word reading outcome measure at a later timepoint. Included studies reported correlation coefficients to quantify relationships between DAs and SAs and/or DAs and word reading outcome measures.
iv. Articles published in English, French or Spanish, or those written in another language but with full text translations were included. No exclusions were made based on setting.

### Search Strategy and Information Sources

An initial search was carried out in March 2022 on the following 5 databases: MEDLINE, Embase, CINAHL (Cumulative Index to Nursing and Allied Health Literature), PsycINFO and ERIC (Education Resources Information Center), using two concepts “dynamic assessment” and “literacy” and their associated keywords in titles and abstracts. Synonyms for dynamic assessment included (dynamic test* OR screen* OR tool* OR task* OR measur*) OR (learning potential assess* OR screen* OR test* OR tool* OR task OR measur*), OR (response to intervention). Associated keywords for literacy included phonem* OR phonolog* OR phonic* OR (sound* blend* OR segment* OR manipulat* OR substitut* OR delet*), OR (letter* OR alphabet* knowledge or principle) OR read OR reading OR write OR writing OR spell OR spelling OR decode OR decoding. No filters were used in the search process. This search strategy was developed with support from the University of Toronto librarian. Following the initial database search, in June 2022 an additional synonym for “dynamic assessment” was added – computerized adaptive testing. This search term (comput* adapt* test*) was rerun on the same databases with all synonyms for the key word “literacy” and results of this search were screened. Computerized adaptive tests (CAT) were not identified as a method/synonym of DA in the initial preliminary searches but were determined to be necessary to search following identification of a study that utilized a dynamic CAT approach to reading assessment. For a list of search terms used in each database see Figure 1 and Figure 2 in Appendix 1.

Next, a search was performed in 3 preprint repositories, MedRxiv, EdArxiv and PsyArxiv. The same two concepts “dynamic assessment” and “literacy” were used to conduct this search. Following the database and preprint repository search, the first author and second author began forward searching of included articles on Google Scholar. The “cited by” function was used to identify articles that had cited the included/relevant studies identified from the database and preprint search. Subsequently, an ancestral search of the included articles was then conducted. The reference lists of the included articles were reviewed by one of ten coders or the first author and crosschecked with the included article list to determine if there were any articles of interest that had not been identified in the database, preprint, or Google Scholar search. Finally, requests for unpublished data or studies were posted to lab and researcher social media accounts and sent out on two occasions to relevant mailing lists, and to labs across Canada, the United States and Europe conducting early literacy research. An updated search of databases and preprint repositories was conducted in December 2022 to identify new relevant articles.

At each stage of screening, articles were rated by two independent reviewers, either the first author or one of ten trained research assistants (RAs). In the title/abstract stage reviewers voted whether an article was relevant or irrelevant, in the full-text eligibility screening, reviewers voted to include or exclude a study based on its’ characteristics. In all phases, disagreements were resolved by the first author. Given that 11 reviewers participated in the article identification process, there were many unique pairings of raters (i.e., 66 pairings at the title/abstract stage). Consequently, calculation and reporting of Cohen’s Kappa coefficients for all reviewer pairings was not meaningful. Rather, the average Kappa coefficient for all rater pairs was calculated to be 0.29 (90% proportionate agreement) for the title abstract screening and 0.39 (76% proportionate agreement for the full text review screening, which can both be characterized as fair (Cohen, 1960; McHugh, 2012).

### Data Collection Process

Following identification of relevant articles, data was extracted using a template generated on Covidence. The same team who completed the title/abstract and full text screening performed the extraction. All coders received a training session led by the first author prior to extraction. All articles were extracted by two reviewers. Conflicts and consensus were completed by the first author. For each study, the following data points were extracted:

### Data Items

#### General Information

Study title, journal, year of publication, the DOI, author names and institutional affiliations, the country in which the study was conducted, and whether the project received any funding or reported any conflicts of interest.

#### Study Type

Studies were coded as **c**ross-sectional or longitudinal. Longitudinal studies that also included a cross-sectional correlation between SA and DA measures at a single timepoint and a correlation between DA and a reading OM across two timepoints were counted as both cross-sectional and longitudinal.

#### Participants

For each study, the total number of participants, the percentage of males and the mean age and grade at the study outset were coded. Age and grade are not consistent across countries so both data points were required. The reading status (typically developing, at-risk or diagnosed with a reading difficulty- or any combination of these groups), and the language status (monolingual, bilingual, or a combination of both) of the study participants was coded, along with which language(s) were reported to be spoken by the participants.

### Measures

#### Dynamic Assessment(s)

DAs in this review are defined as any assessment which provided explicit or implicit teaching, training, feedback on performance or prompting in the context of the assessment. Coders extracted the name of the DA if one was provided, the word reading skill(s) that the DA evaluated (either PA, SSK or decoding, or a combination of two or three of these skills) and a brief description of the specific task used to evaluate the literacy skill (e.g., PA-phoneme segmentation, or SSK-letter-sound knowledge of the English alphabet). If more than one task was used to evaluate a skill, as was often the case, coders listed all tasks. These data points permit comparison of the concurrent and predictive validity of each construct of word reading DA.^1^

#### Static Assessment(s) and Word Reading Outcome Measures (OMs)

SAs and OMs in this review are any assessments which evaluate a skill using a binary correct/incorrect response scoring system, and which are characterized by the absence of feedback, prompting or training and teaching components in the assessment. SAs in this review are tests that are conducted at the same time point (concurrently) as the DA, while OMs are tests that are conducted at a later time point and used to investigate predictive validity of DA. Both SAs and OMs in this review can be norm-referenced tests (e.g., CTOPP), criterion-referenced tests (e.g., the Dynamic Indicators of Basic Literacy Skills) or researcher developed tools. When extracting information related to SAs, coders indicated the name and type of any assessments used (e.g., CTOPP, norm-referenced), the word reading skills evaluated (PA, SSK, Decoding or a combination) and the specific tasks used to evaluate these skills (e.g., PA-phoneme blending, SSK-novel symbol-sound knowledge). Regarding outcome measures, coders identified the name, if any, of the outcome measure used (e.g., WRMT-R NU) and the specific subtests of the measure (e.g., Word Attack) and the skill evaluated (e.g., nonword reading accuracy).

#### Effect Sizes

The correlation coefficients representing the relationships between the DAs and SAs, and/or the DAs and OMs were extracted. If a study reported multiple correlations between a DA and an SA (e.g., a DA of PA that utilized multiple PA tasks, and a SA evaluating PA that also employed multiple PA tasks), or a DA and an OM, coders were instructed to extract all relevant correlations coefficients at this stage. Following review of all extracted coefficients, the authors created a set of decision rules for choosing a single correlation coefficient to represent the relationship between the DA and SA, or the DA and the OM for each analysis, to ensure that the synthesis did not violate the assumption of independence. These decision rules for selecting a single effect size were made based on which measure was most consistently used across studies. For example, word reading accuracy was reported in the majority of included study as an outcome measure, and as such it was identified as the primary outcome measure for estimating predictive validity. Effect sizes between DA and WR accuracy were selected over those that were less commonly observed, like non-word reading, passage reading, or word reading fluency. Similarly, because most studies examined Kindergarten aged students at time point 1, and Grade 1 students at time point 2, effect sizes representing the association between a DA in kindergarten and an OM in grade 1 were favoured over those representing less commonly observed time points such as preschool and grade 2.

In instances where when one measure was not more common than all others, decision rules grounded in literacy theory were used. For example, there was not a single most used PA task, and so, when possible complex PA tasks (e.g., manipulation) were preferred over simple phonemic awareness tasks (e.g., blending), and smaller unit tasks (e.g., phoneme level) were preferred over larger unit phonological awareness tasks (syllable level tasks) as these complex, smaller grain, tasks have consistently been linked to later decoding success (Høien et al., 1995). In terms of SSK tasks-those which required a child to name a make a connection between a symbol and a sound were preferred over those that required mere naming of the symbol. This is because this skill more closely approximates the construct of the alphabetic principle, which research has determined is an excellent predictor of later reading ability (Ehri, 1998). Finally, when choosing decoding tasks – single real word, untimed, decoding tasks were prioritized over timed, nonword or passage level decoding tasks as this best represents the construct of decoding as it is defined in this review. The coefficients representing the effect sizes for concurrent and predictive validity are presented in Tables 5 and 6, respectively.

### Quality Appraisal Assessment

Two trained independent reviewers assessed the included studies using a modified and combined version of two quality assessment tools for (i) cross sectional design and (ii) diagnostic accuracy studies from the Johanna Briggs Institute (Moola et al., 2020). Studies were evaluated on the following five domains (i) participant selection, (ii) index/dynamic assessment, (iii) reference/static assessments and/or outcome measures, (iv) flow and timing of the study, (v) statistical analysis. Please refer to Table 3 in Appendix 2 for the full list of questions and ratings for each study.

Regarding participants, reviewers rated whether the participant sample was adequately described in terms of age, sex breakdown, language and reading status and demographic characteristics. Reviewers evaluated the DA domain by rating whether it was described with sufficient detail in terms of the skills evaluated, the format of the test, the prompting and scoring used, and administration process. They also recorded whether the task used to evaluate the word reading skill(s) in the DA was developmentally appropriate for the population. When assessing the domain related to the reference standards (SAs and OMs) reviewers evaluated whether the studies employed developmentally appropriate tools for evaluating word reading skills or word reading outcomes. They also rated whether psychometric properties of the reference measures used were reported. In evaluating flow and timing, reviewers noted whether all participants were included in the analyses and whether authors explained and accounted for reasons for loss to follow up and attrition if necessary. Finally, coders assessed whether appropriate statistical analyses were used to draw conclusions about study findings.

In summary, this yielded 8 items across the 5 domains to be rated. Items relating to participants, flow and timing and statistical analyses were weighted one point, while items pertaining to the index test (DA) and the reference tests (SA and OMs) were weighted two points given their relative importance in addressing the review objectives. Following ratings by two reviewers, conflicts were resolved by the first author, and studies were ranked as either low quality (0-33%), medium quality (34-67%) or high quality (68-100%). Only studies rated as medium and high quality were included in the analyses. Please refer to Appendix 2 Table 3 for the questions and study ratings.

### Analyses

We used a random effects model for our meta-analyses (Borenstein et al., 2010), and included subgroup analyses by DA type (PA, SSK and decoding), language status (monolingual, bilingual and mixed) and reading status (typically developing, at-risk/diagnosed with reading difficulty and mixed).

The ‘metacor’ package (Laliberté, 2019) in R studio (R Core Team, 2021) was used to conduct a Fisher Z transformation of Pearson’s correlation coefficients into Z scale scores (Corey et al., 1998; Silver & Dunlap, 1987). Following the transformation, a weighted average of these values was then calculated and transformed back to Pearson r correlation coefficients with accompanying p values for interpretation. To provide a robust picture of the degree heterogeneity between studies, Q, *I*^2^ and τ^2^ heterogeneity statistics were calculated (Borenstein et al., 2017; Higgins & Thompson, 2002). To evaluate which studies contributed most to overall heterogeneity, Baujat plots were generated and are presented in Figure 12 and 13 in Appendix 3 (Baujat et al., 2002). To examine the potential risk of publication bias, funnel plots for both the concurrent and predictive validity analyses were generated in R studio (R Core Team, 2021). Egger’s regression test, a test of significance, was conducted to determine objectively whether funnel plot asymmetry was present (Egger et al., 1997).

## Results

### Study Selection

The process of study identification is visualized in Figure 3 in the PRISMA flowchart below (Page et al., 2021). The database searches produced 4,626 articles which were uploaded to Covidence. The software automatically detected and removed 1408 duplicate articles leaving 3218 titles and abstracts for review. Of these articles, 138 were reviewed at the level of full text, and 23 articles were identified for inclusion. Next, the 876 articles identified from the preprint repositories searches were uploaded. There were 0 duplicates. Following title/abstract screening, 3 articles were reviewed as full texts, and 1 article was included for analysis. An additional 1,351 articles were identified via forward Google Scholar searching of the 24 already included articles. Over three rounds of iterative searching, 17 relevant articles were identified, of which 11 were excluded and 7 included. Subsequently, an ancestral search of the 31 identified articles was completed. This yielded 15 potential articles reviewed at the full text level. One additional study was deemed relevant for inclusion. Finally, the callout to social media, mailing lists and labs was made. Two authors contacted the first author to share 4 papers. 3 papers were reviewed at the full text level and 0 were included. In summary, 32 papers including a total of 34 studies were identified for inclusion in the systematic review and meta-analysis. The process of study identification is visualized in the PRISMA flowchart below (Page et al., 2021). Reasons exclusion are also reported.

**Figure 3.**
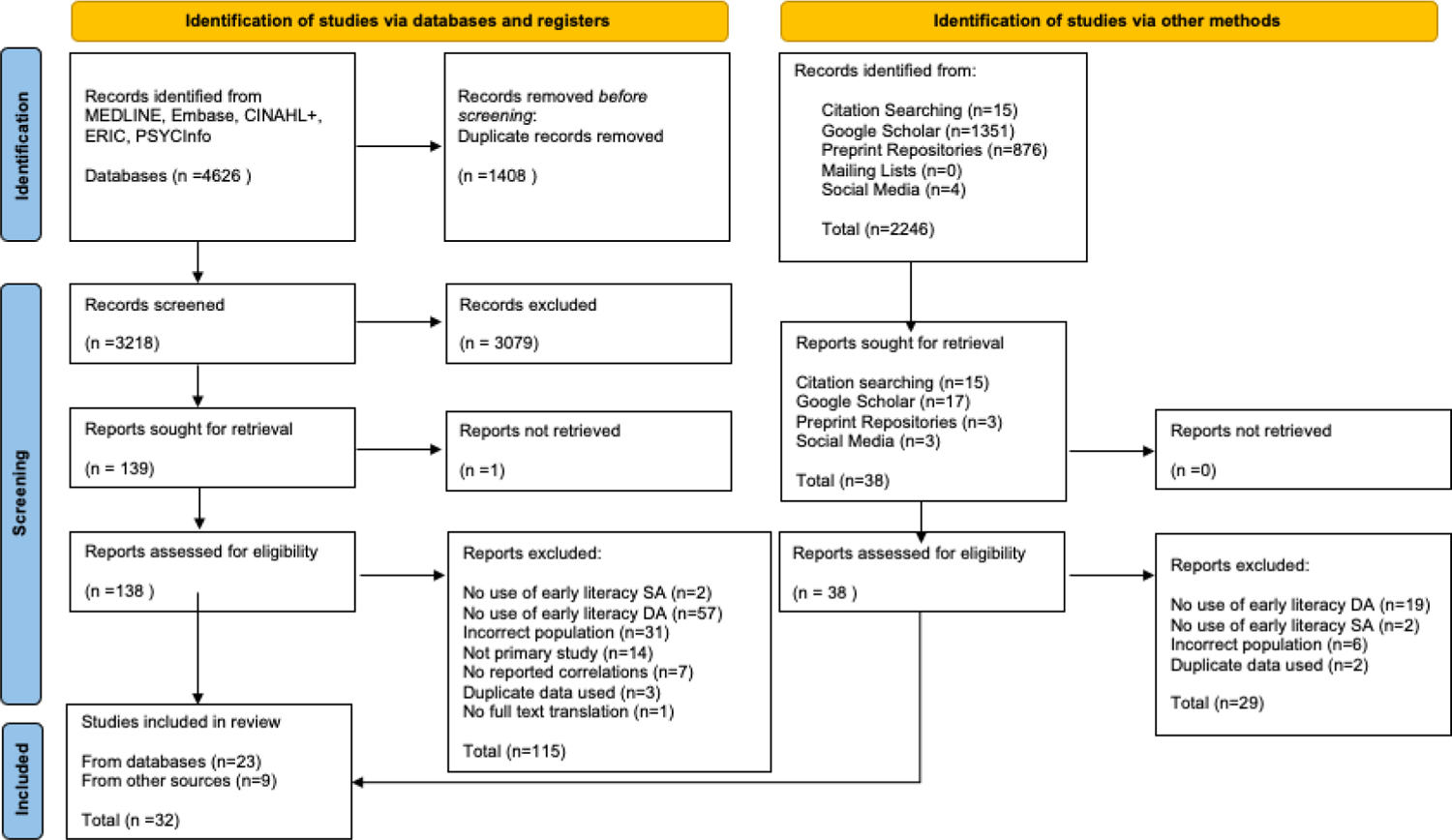
PRISMA Flowchart of Literature Search

### Study Characteristics

#### Age and Sex

A total of 6791 participants were included across 34 studies in 32 included articles. The mean age across participants was 5;8. The most common grade tested at a single time point (for cross-sectional studies) or at timepoint 1 (for longitudinal studies) was Kindergarten, and the most common for follow-up grade was approximately one year later in grade 1 (n=9) while the second most common follow-up age was later in the kindergarten year (n=6). Across studies, the mean % of males were 50.92%. Mean age, % of males included and grade at start was not reported in all included studies. Table 1 below lists the breakdown of mean age and % males across subgroups stratified by language and reading status.

**Table 1.** Participant Characteristics

|  | N | Mean start age* | Mean % males** |
| --- | --- | --- | --- |
| **Total**<br>(min-max) | 6696<br>(9-1988) | 5;8<br>(4;1 – 9;9) | 50.92%<br>(41%-67%) |
| *Language Status* |  |  |  |
| Monolinguals | 3812 | 6;5 | 51.1% |
| Bilinguals | 231 | 5;3 | 49.1% |
| Mixed language | 2653 | 5;2 | 48.03% |
| *Reading Status* |  |  |  |
| Typically developing | 3445 | 5;2 | 50.85% |
| At-risk | 348 | 7;7 | 52.4% |
| Mixed reading | 2903 | 6;4 | 49.8% |

*Note.* N= Number of participants
\* 23/34 studies reported mean age of participants at study onset
\*\*26/34 studies reported mean % males in their participant sample

#### Languages Spoken and Language Status

The majority of studies were conducted in the United States (n=17), followed by the Netherlands (n=3), Denmark (n=2), China (n=2), England (n=2), Germany (n=2), Belgium (n=1), Finland (n=1), Singapore (n=1), Spain (n=1) and Taiwan (n=1). Of the 34 studies, 24 included monolinguals only. The languages spoken by the monolinguals included English (n=15), Danish (n=2), Dutch (n=2), Mandarin (n=2), Finnish (n=1), Spanish (n=1) and German (n=1). Six studies included both monolingual and bilingual populations in their analyses. Of those 6, only 1 specified the languages spoken by both the mono and bilinguals in their study (English monolinguals and Spanish/English bilinguals). All 5 other studies did not provide linguistic details about the included bilingual groups but did indicate that the monolinguals spoke Danish (n=2), German (n=1), English (n=1), or English, Swedish or Norwegian (n=1). Finally, five studies included only bilingual groups. Of those, three were conducted with Spanish/English bilinguals, 1 with Mandarin/English bilinguals and 1 with English/Chinese, Malay, or other bilinguals. Please see Table 2 below for further information about the language status and languages spoken by participants in included studies.

**Table 2.**
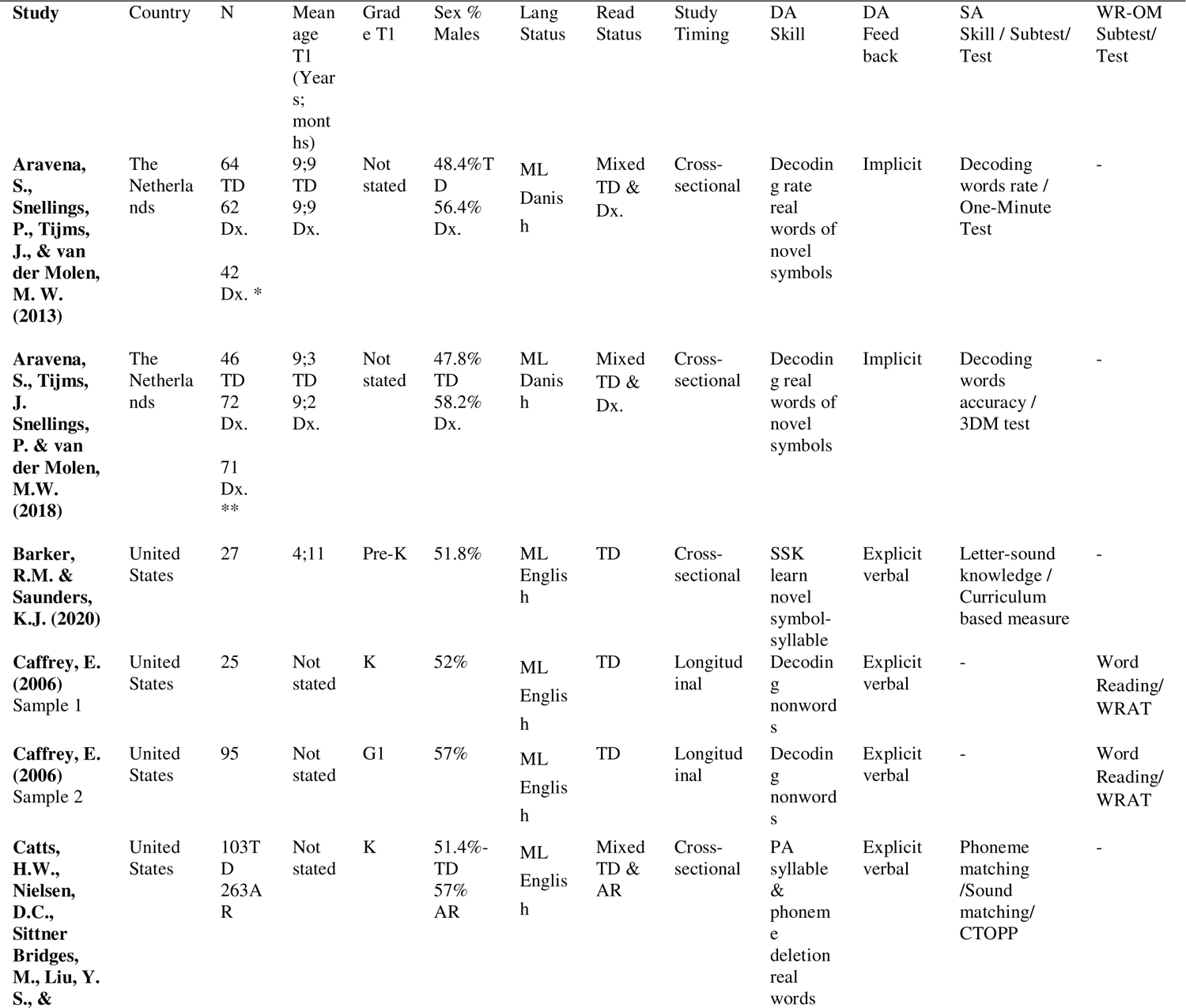
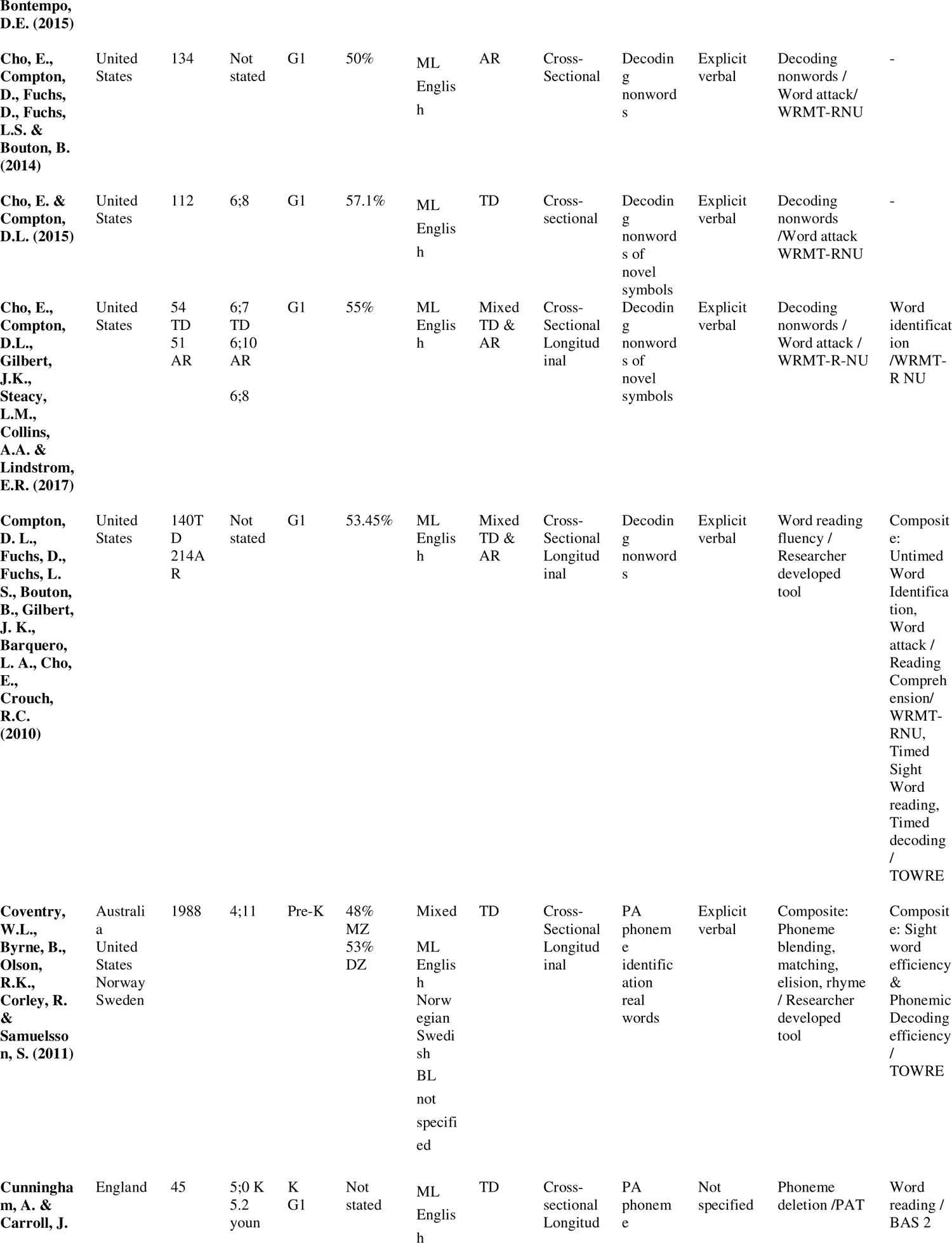
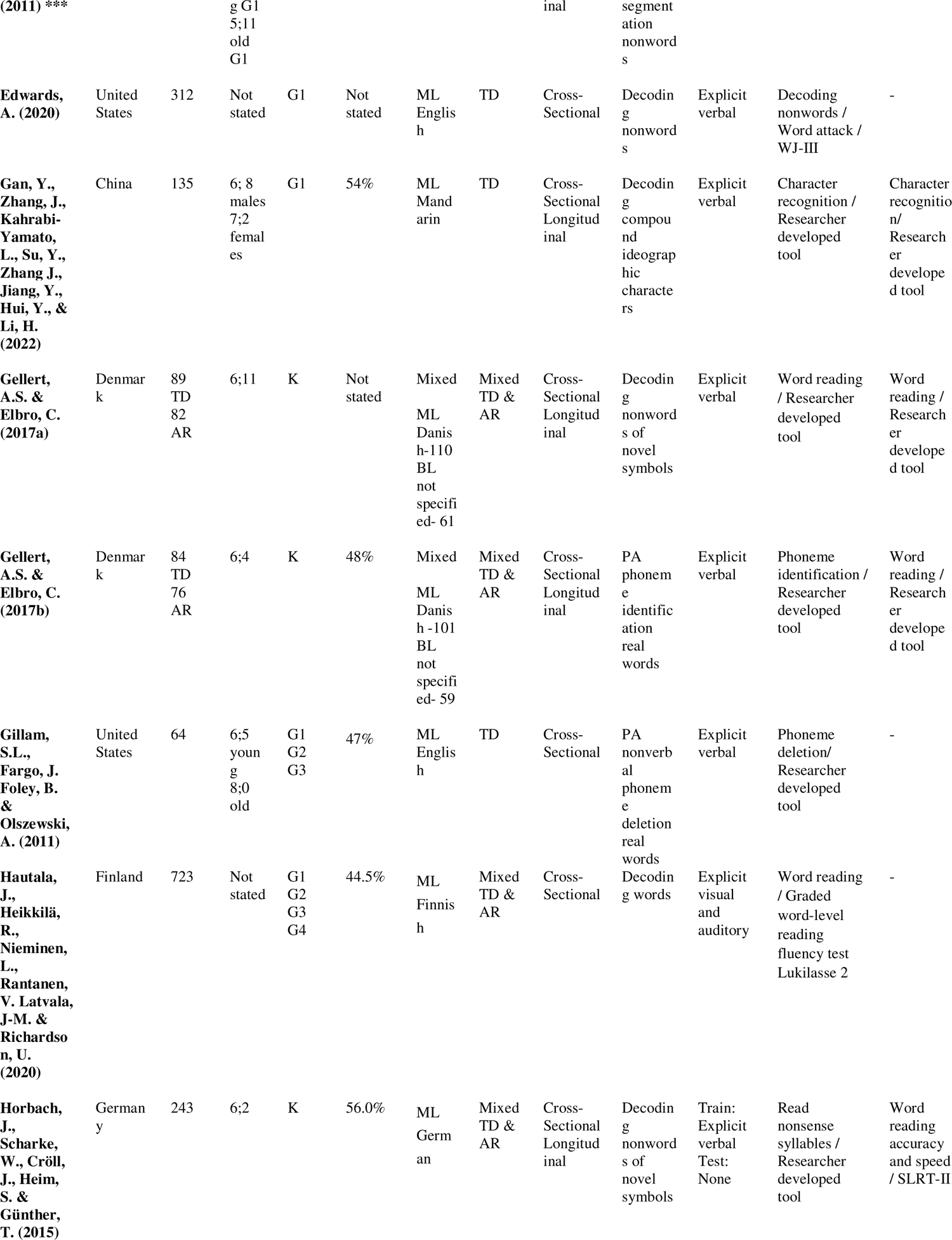
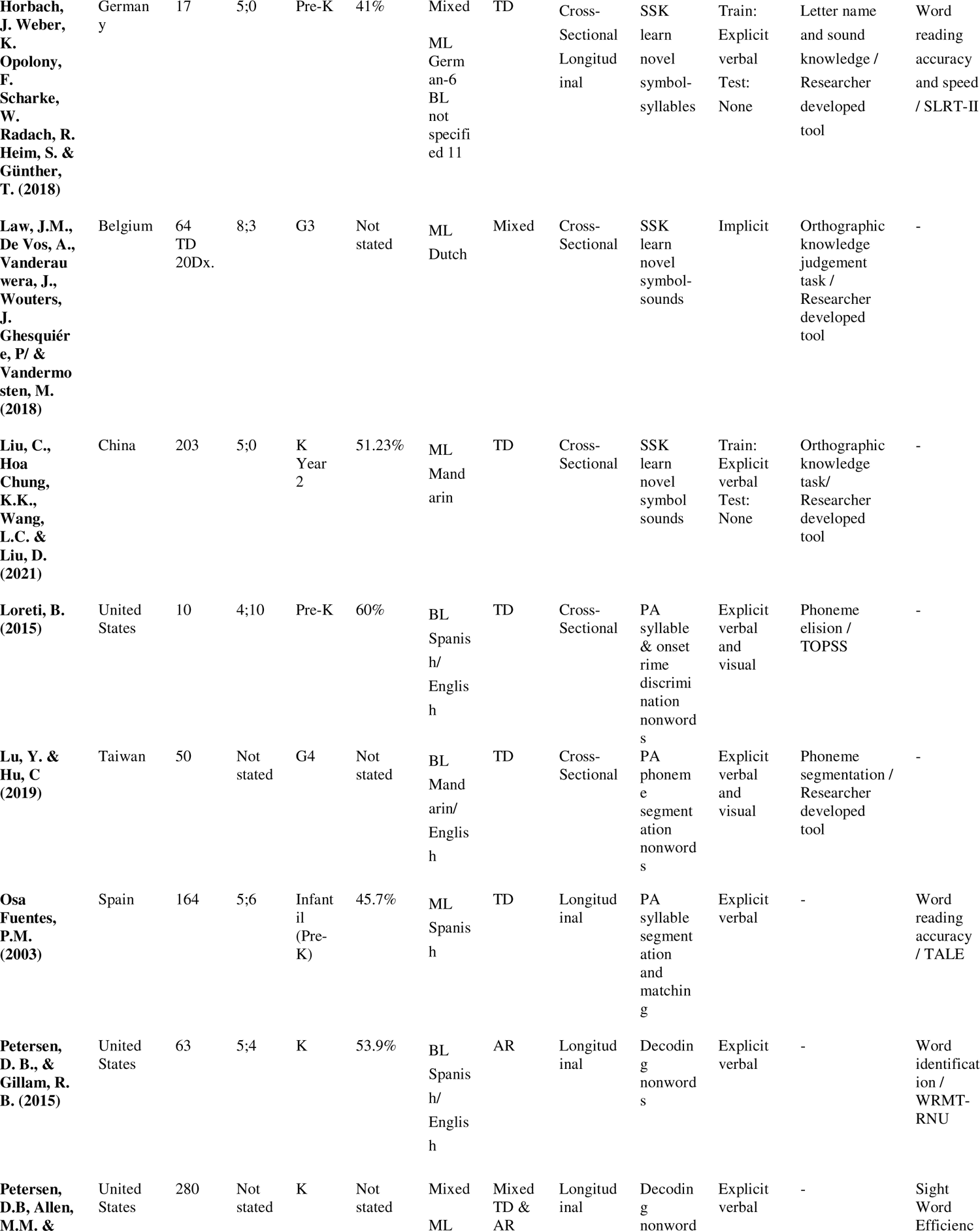
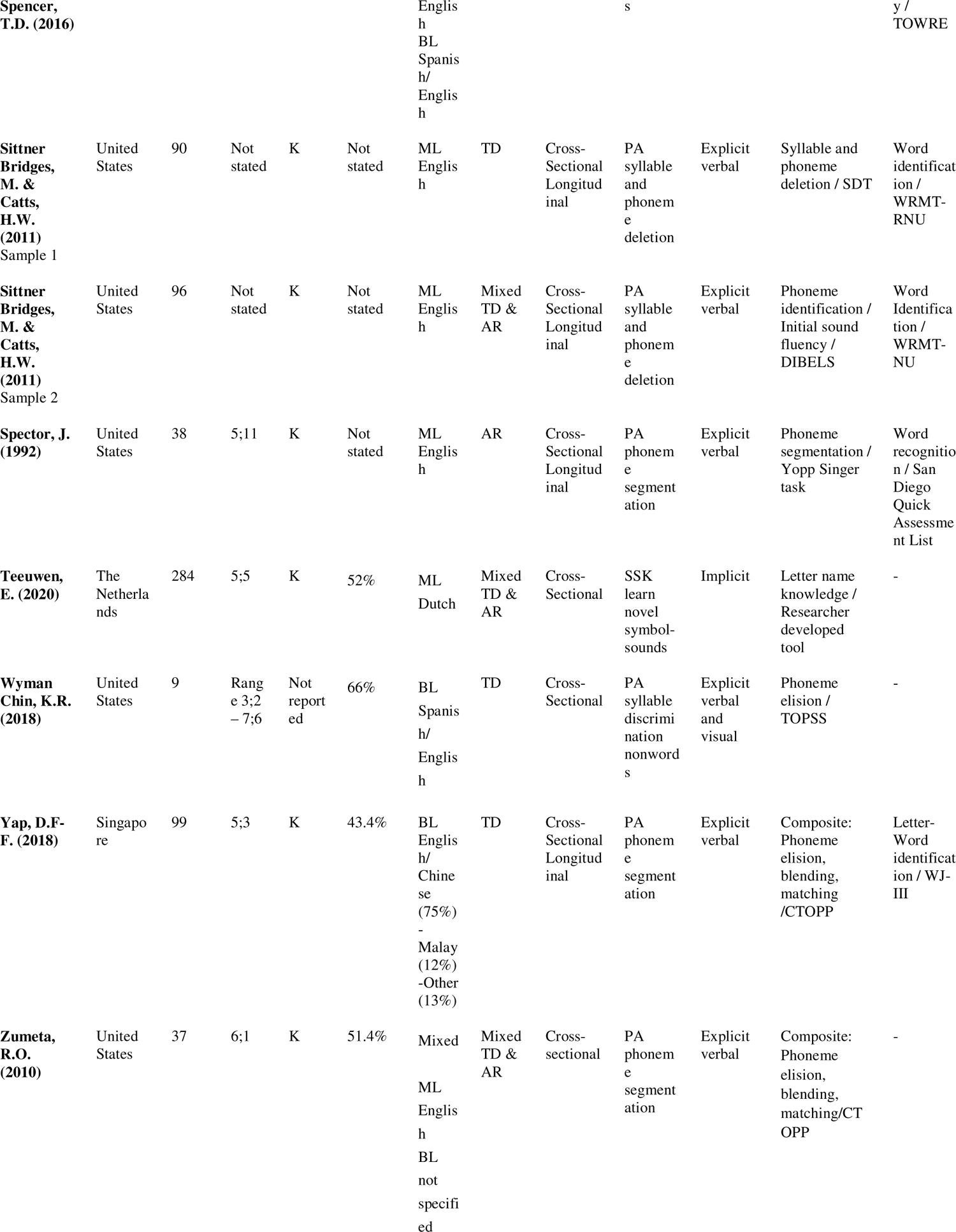
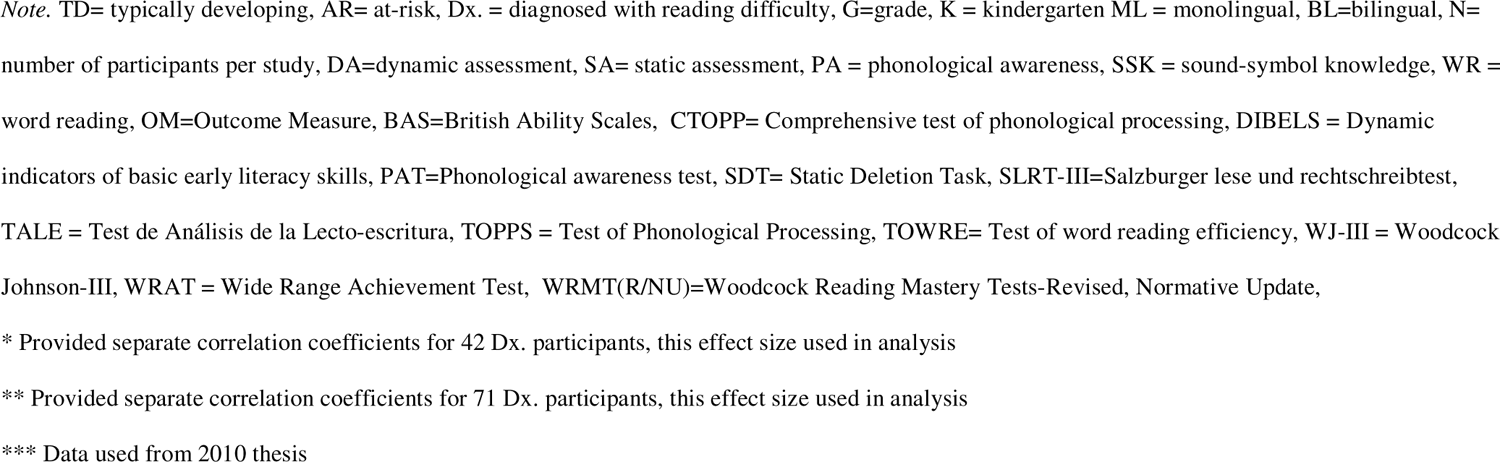
Country, Number of Participants, Mean Age, Grade, % Males, Language Status, Reading Status, Study Design, Type and Characteristics of DAs, SAs and WR Outcome Measures of Included Studies

#### Reading Status

Seventeen of the 34 studies included exclusively participants who were typically developing. Two studies conducted their correlational analyses separately for participants diagnosed with dyslexia, and 3 exclusively included participants at-risk for reading difficulty. Given the limited number of studies conducted with exclusively at-risk children or those diagnosed with difficulty, this group was merged. Finally, 12 studies included a mix of typically developing and at-risk groups in their analyses. Table 2 provides more information about participants’ reading status in included studies.

#### Dynamic Assessments

Fourteen of the included studies evaluated phonological awareness via syllable or phoneme identification (n=5), syllable or phoneme deletion (n=4) and segmentation (n=5) tasks. Five DAs evaluated sound-symbol knowledge via novel symbol-sound or symbol-syllable learning tasks. Fifteen DAs evaluated decoding via either nonwords (n=11) or real words (n=4) and using novel symbols (n=7) or known letters (n=8). Four studies used implicit feedback via a game in their DA, while the remaining 30 employed explicit verbal and/or visual feedback. Table 2 provides additional information about characteristics of DAs in the included studies.

#### Static Assessments and Word Reading Outcome Measures

Of the 29 studies that reported a correlation between a DA and an SA, the majority used a norm-referenced tool as the SA (n=15), followed by researcher developed tools (n=12) and standardized screening tools (n=2). The majority of the 18 longitudinal studies included in this review used a norm-referenced tool at follow-up to evaluate word reading abilities (n=14). Fewer used researcher developed tools (n=3), or standardized screening tools (n=1). The most common word reading outcome measure was the Word Identification subtest (n=6) from the Woodcock Reading Mastery Test-Revised, Normative-Update. Table 2 below provides additional information regarding characteristics of SAs and word reading outcome measures in the included studies (e.g., names of tests and subtests).

### Quality Appraisal

All 32 articles were appraised using the modified Johanna Briggs Institute quality appraisal tool (Moola et al., 2020). The average rating was 10.6/12 or 88%. The most common reason for deduction of points was inadequate description of participants (either based on age, sex, language or reading status). No studies were rated as low quality (0-33%), 1 article (Teeuwen, 2020), was rated as medium quality (33-67%), and the remaining 31 articles were rated as high quality (68-100%). No studies were excluded from analysis based on their quality appraisal rating. See Appendix 2 Table 3 for quality appraisal ratings of included articles.

### Research Question 1A: Do dynamic assessments of word reading skills (phonological awareness (PA), sound-symbol knowledge (SSK), and decoding), demonstrate concurrent validity with static assessments of word reading skills (PA, SSK, decoding) across all populations?

As Table 5 indicates, 28 articles including 29 studies reported correlations between a DA and an SA of an equivalent construct. Thirteen studies examined PA, 5 examined SSK, and 10 examined decoding. Correlations representing the relationship between DAs and equivalent SAs of word reading skills are reported in Table 5 below.

**Table 5.**
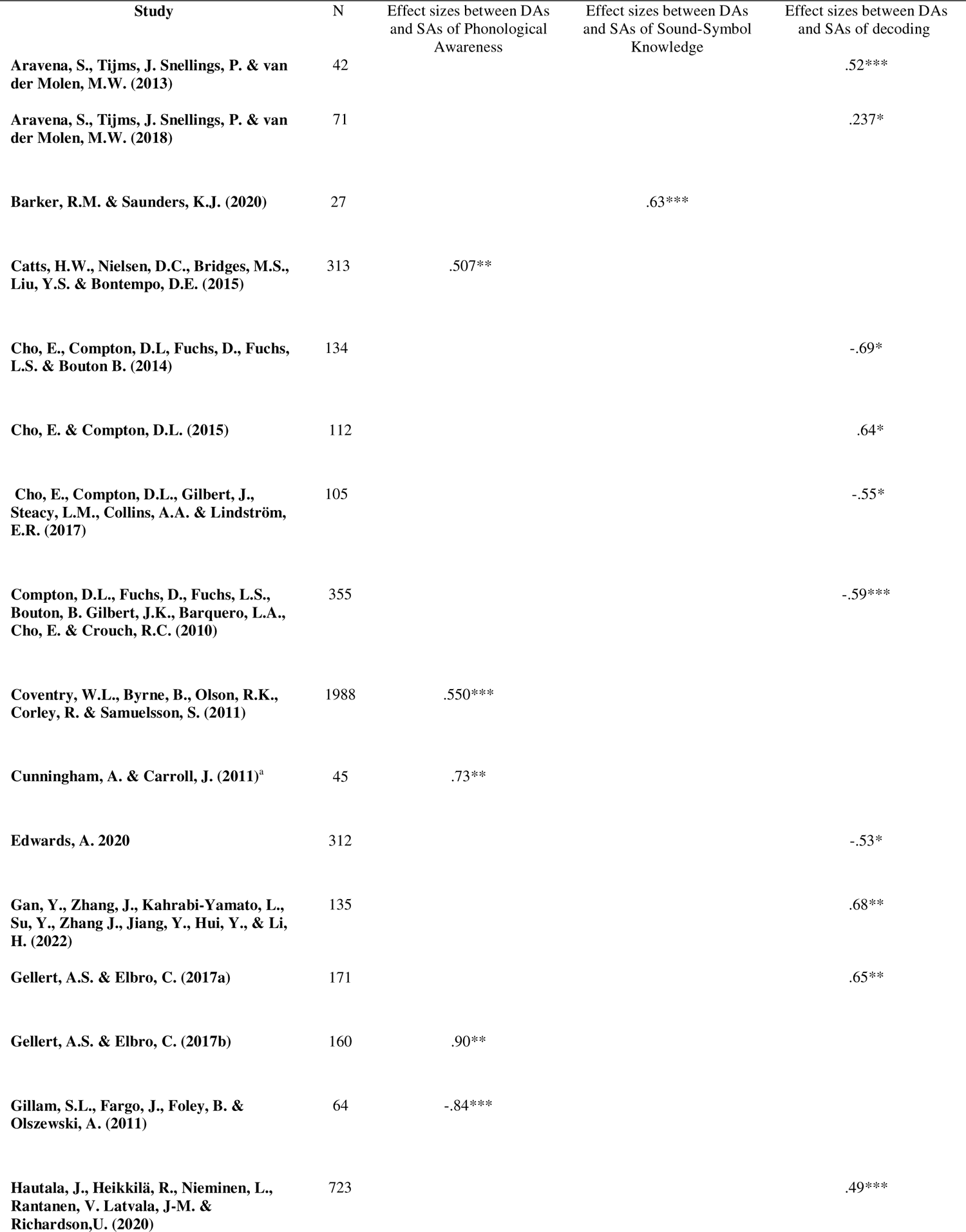
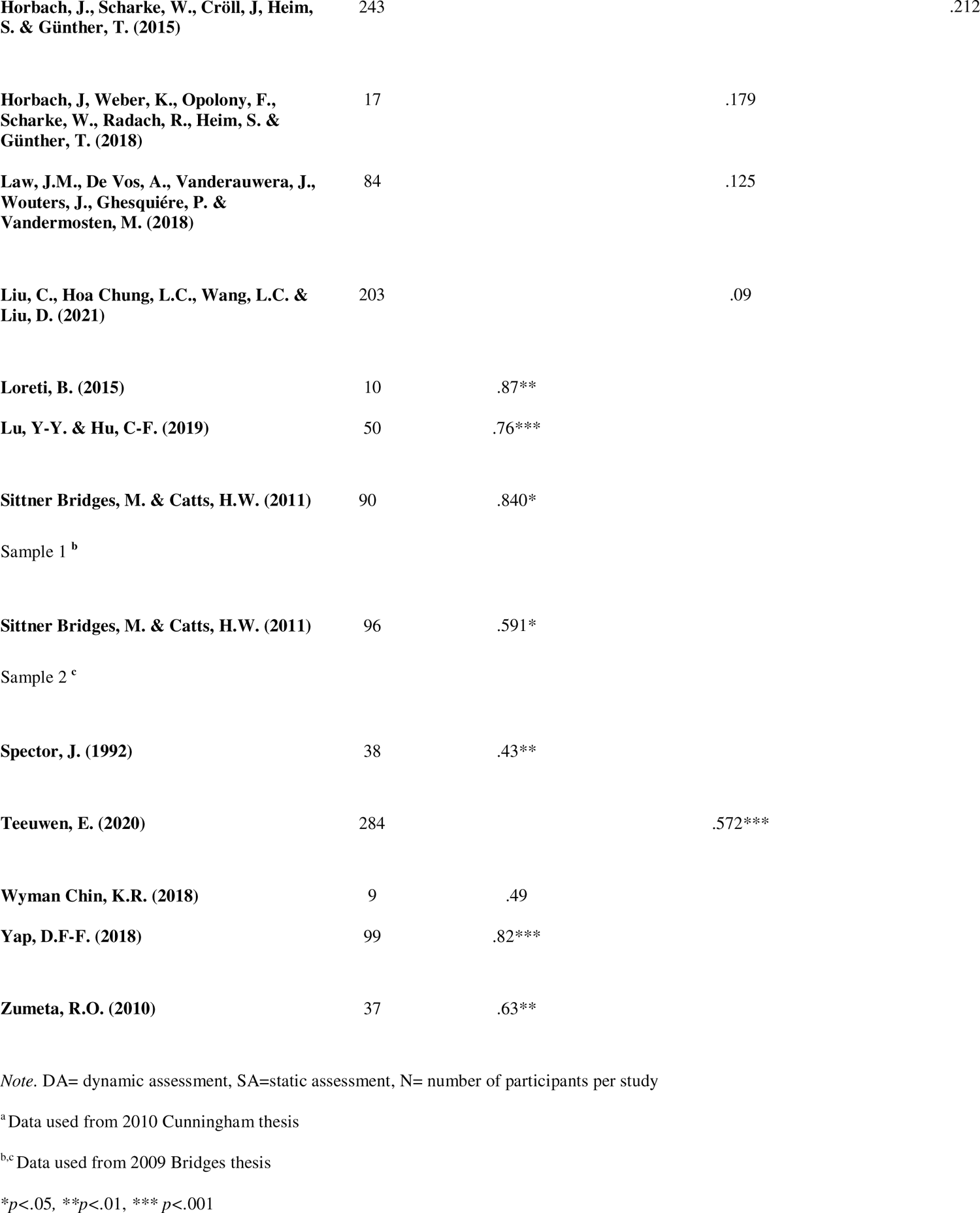
Effect Sizes Representing Concurrent Validity Between Dynamic Assessments and Static Assessments of Word Reading Skills

### Meta-analysis of Concurrent Effect Sizes

The effect sizes from these twenty-nine studies in Table 5 were included in the analyses. Results of the random-effects meta-analysis examining the concurrent validity between DAs and SAs of word reading skills are reported in Figure 4. The overall mean effect size is large (r=0.60, 95%CI = [0.51-0.68]) suggesting that overall DAs are strongly correlated with SAs. Given the significant heterogeneity, the moderator of DA type was included and found to be significant (Q=14.32, df=2, p<0.01). Results of the subgroup analysis are also displayed in Figure 4. Outcomes of this mixed effects model suggest that there is a significant variation between subgroups of DA type (PA, SSK, and decoding) and their concurrent validity with SAs of equivalent constructs. Overall mean effect sizes representing the relationship between DAs of PA and decoding, and their SA counterparts are strong, while the effect size for DAs of SSK is moderate. Furthermore, the prediction intervals for the SSK and decoding subgroups cross 0 indicating that future relevant studies examining relationships between SAs and DAs of SSK and decoding may demonstrate a negative correlation. Importantly, the prediction interval for the PA and decoding subgroups do not cross 0, suggesting that future studies are likely to report positive correlations between these DAs and SAs (Harrer et al., 2021; IntHout et al., 2016). Therefore, the results indicate that DAs of PA skills are most likely to be valid alternatives to traditional SAs, followed by DAs of decoding, which demonstrate strong concurrent correlations a slightly larger prediction interval, and finally DAs of SSK which are associated with weaker overall mean effect sizes and the largest prediction interval. However, it is also important to note that the test for within group heterogeneity in the mixed effects model was still found to be significant, even with DA type as a moderator (Q=307.65, df=25=6, p<0.01), which indicates that there are likely other moderators beyond DA type that are impacting heterogeneity.

**Figure 4.**
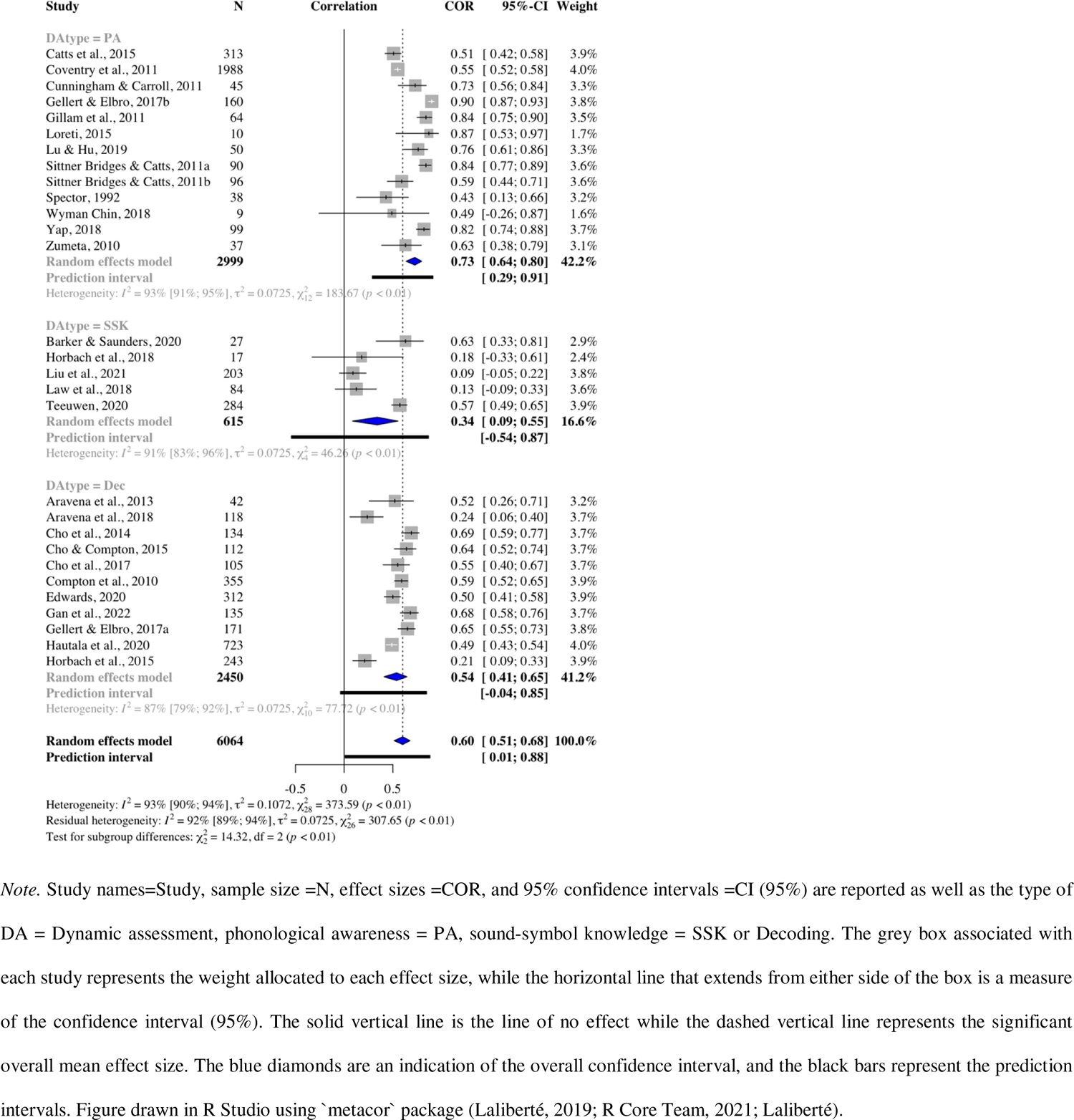
Forest Plot of Random Effects Meta-Analysis Examining the Concurrent Validity Between Dynamic and Static Assessments of Word Reading Skills

### Research Question 1B: Do dynamic assessments of word reading skills demonstrate predictive validity with reading outcome measures (single word reading) across all populations?

Sixteen articles including 18 studies reported correlations between a DA and a later word reading outcome measure. Eight examined PA, 1 examined SSK, and 9 examined decoding. Table 6 below shows correlations between each of the three DA skills and word reading outcomes.

**Table 6.**
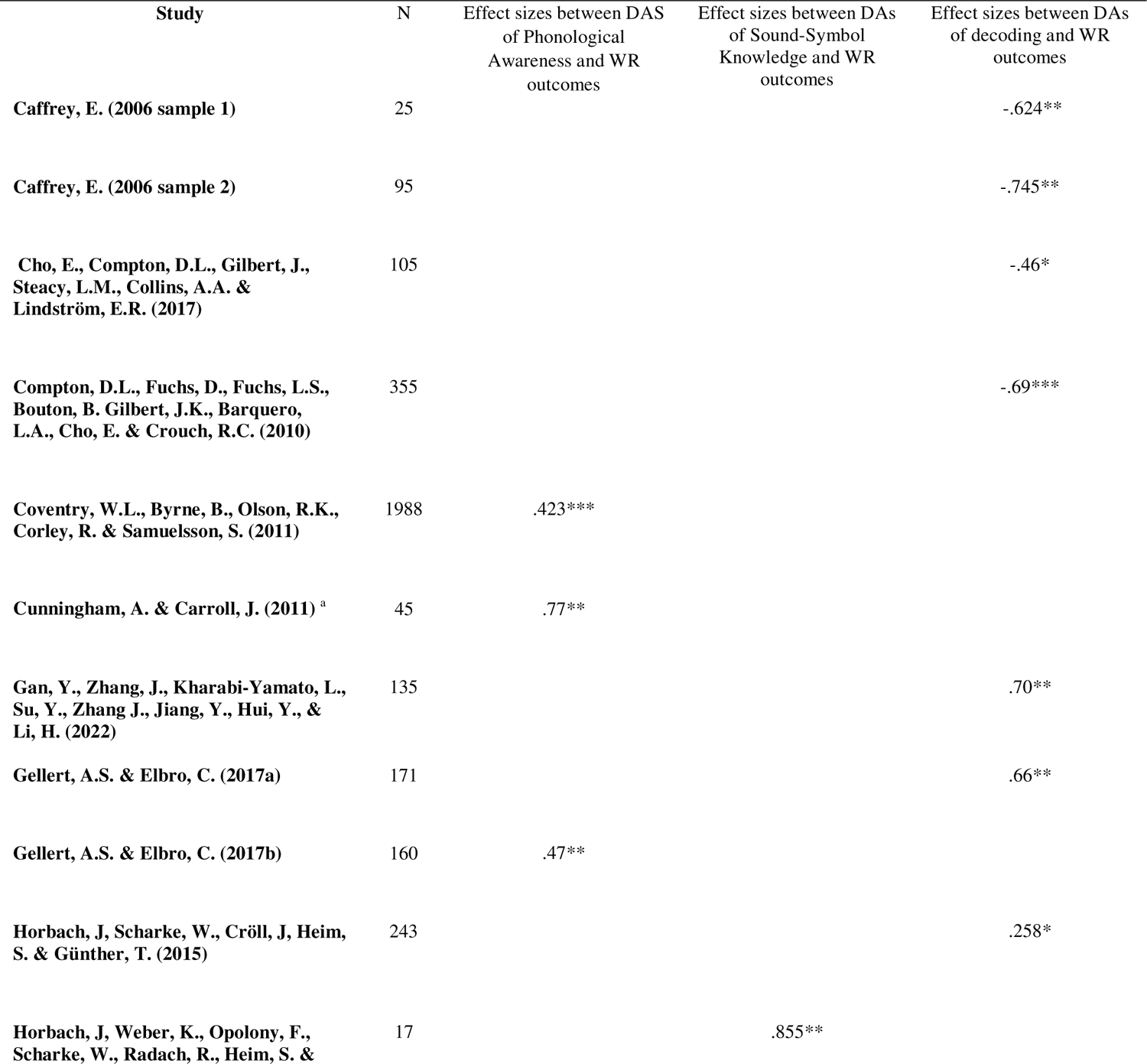
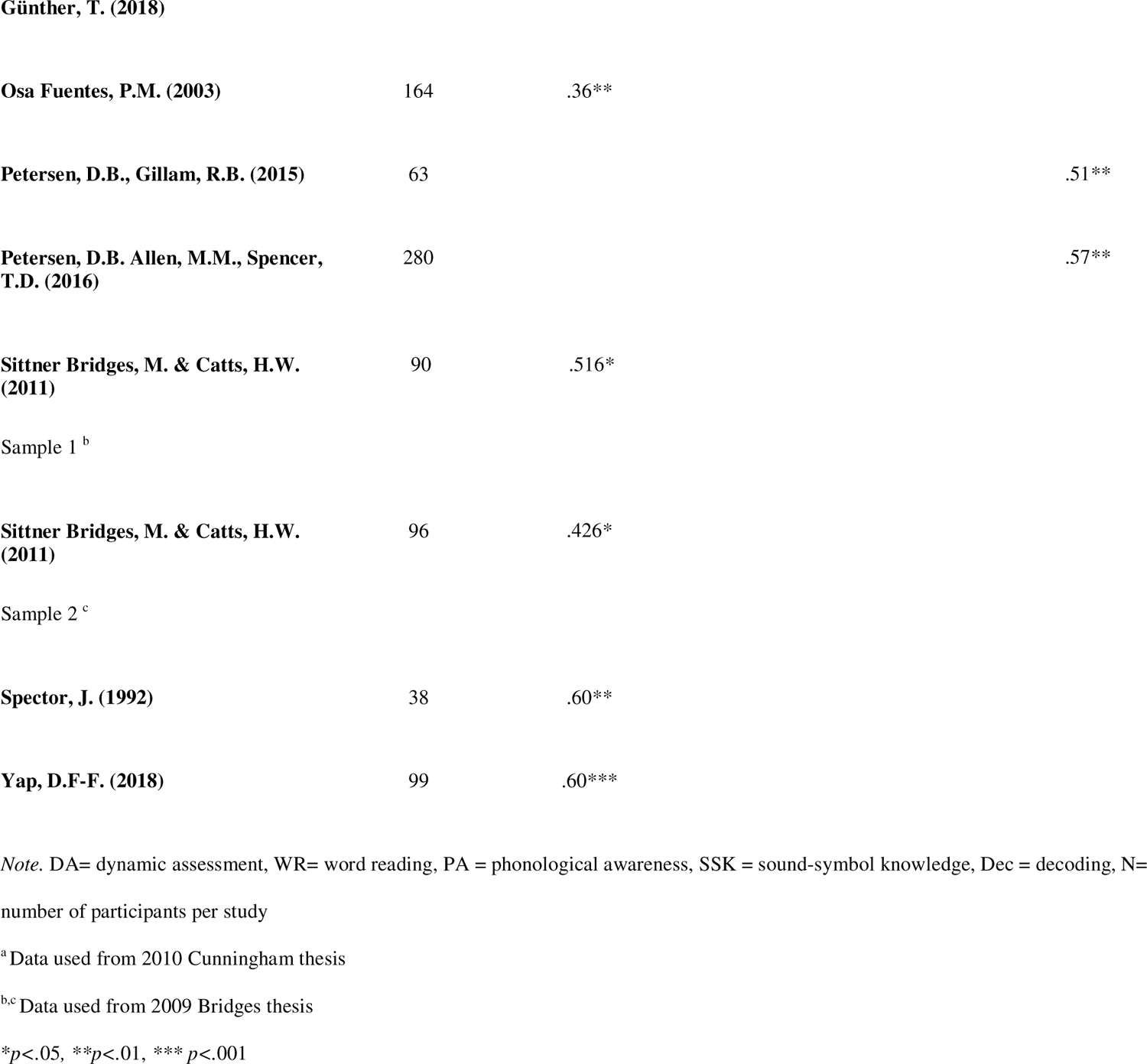
Effect Sizes Representing Predictive Validity Between Dynamic Assessments of Word Reading Skills and Word-reading Outcome Measures

### Meta-Analysis of Predictive Effect Sizes

Results of the random-effects meta-analyses examining the predictive validity of DAs of word reading skills with later word reading outcome measure are presented Figure 5. The overall mean effect size is large (r=0.57, 95%CI = [0.49-0.64]) suggesting that DAs are strongly correlated with word reading outcome measures. Given the significant heterogeneity, the moderator of DA type was included but not found to be significant (Q=4.73, df=2, p=0.09). The results of this mixed effects model, also displayed in Figure 5, indicate that there is not significant variation between subgroups of DA type (PA and decoding) and their predictive validity with tests of word reading outcomes as determined by correlation coefficients. The subgroup of SSK could not be compared due to a lack of studies. The overall prediction interval and those representing the relationship between DAs of PA and decoding and later word reading outcomes do not cross 0, indicating that future studies are likely to indicate positive correlations between DAs of PA and decoding and later word reading outcomes (Harrer et al., 2021; IntHout et al., 2016). The results of this analysis provide suggestive evidence for the predictive validity of DAs of PA and decoding with word reading outcomes. However, again it should be noted that the test for withing group heterogeneity in the mixed effects model was still found to be significant, even with DA type as a moderator (Q=92.52, df=15, p<0.01), which indicates that there are likely other moderators beyond DA type that are impacting heterogeneity.

**Figure 5.**
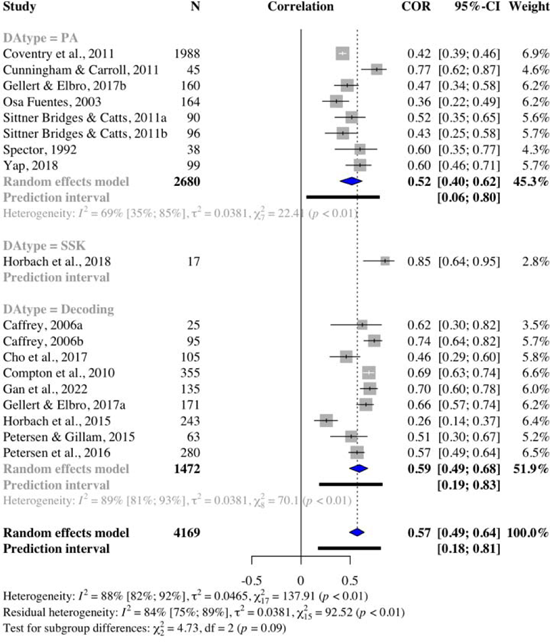
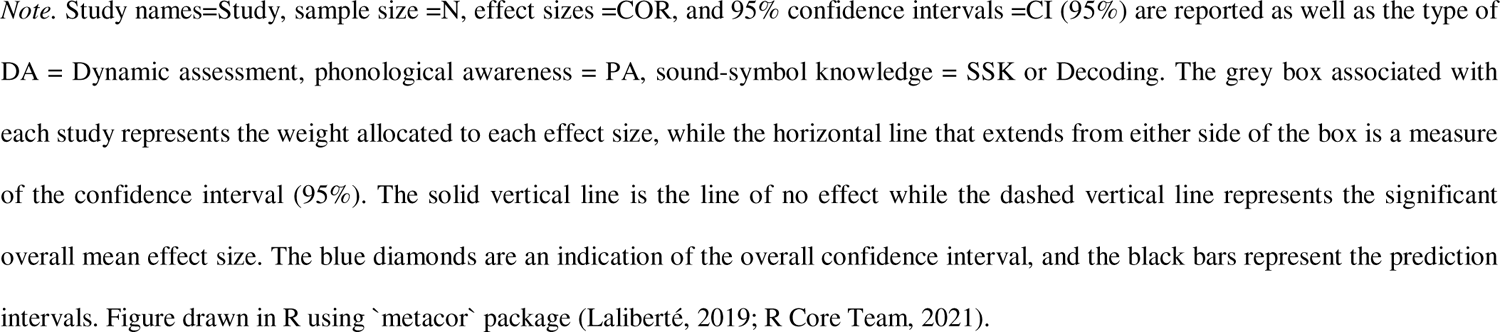
Forest Plot of Random Effects Meta-Analysis Examining the Predictive Validity of Dynamic Assessments of Word Reading Skills with Word Reading Outcome Measures

### Research Question 2A: Do dynamic assessments of word reading skills demonstrate concurrent validity with static assessments of word reading skills, and predictive validity with word reading outcome measures within population groups defined by their language status (monolingual vs. bilingual)?

#### Concurrent validity –Subgroup analysis language status

Of the 29 studies that reported a correlation coefficient between a DA and an SA of an equivalent construct 20 studies included exclusively monolinguals, 4 studies included exclusively bilinguals, and 5 studies included mixed language status populations in their analyses. Table 2 presents further study details. A subgroup analysis by language status was planned a priori. Results are reported below in Table 7. The outcomes of this mixed effects model suggests that there is significant variation between subgroups defined by language status (monolingual, bilingual or mixed mono and bilingual populations) in terms of the concurrent validity of DAs with their equivalent construct SA (Q=6.54, df=2, p=0.04). The effect sizes were largest for bilingual groups, followed by mixed language groups and monolingual groups. Prediction intervals crossed 0 for all three subgroups in this analysis. This outcome runs counter to what was expected. Given that SAs are typically designed for monolinguals, we hypothesized that the overall mean effect size between DAs and SAs would be larger for this group, while we might observe smaller effect sizes between the two for bilinguals who are prone to underperform on SAs. It is possible that this outcome is a result of other factors that cannot be accounted for in a simple subgroup analysis. For example, the limited number of studies in the BL group (n=4) included only typically developing children, while the many studies in the ML group (n=20) included typically developing and at-risk children, as well as those diagnosed with dyslexia. It is possible that this, or other unknown factors, are contributing to this difference.

**Table 7.** Results of Subgroup Analyses by Language Status for Concurrent Validity

|  | Included studies | *g* | 95%CI | *I*<sup>2</sup> | *I*<sup>2</sup> 95%CI | p value heterogeneity | Prediction interval | p value subgroup |
| --- | --- | --- | --- | --- | --- | --- | --- | --- |
| *Language Status* |  |  |  |  |  |  |  | 0.04 |
| **Monolingual** | 20 | 0.54 | 0.44-0.63 | 0.90 | 0.87-0.93 | <0.01 | -0.04 - 0.85 |  |
| **Bilingual** | 4 | 0.78 | 0.58-0.89 | 0.03 | 0.0-0.85 | 0.38 | -0.46 - 0.99 |  |
| **Mixed** | 5 | 0.72 | 0.53-0.84 | 0.97 | 0.95-0.98 | <0.01 | -0.51-0.98 |  |

#### Predictive validity-Subgroup analysis language status

Of the 18 studies that reported a correlation coefficient between a DA and a later word reading outcome measure, 11 included exclusively monolinguals, 2 studies included exclusively bilinguals, and 5 studies included mixed mono and bilingual populations. Table 2 lists further study details. Subgroup analysis by language status was planned a priori and results are reported in Table 8. The outcomes of this mixed effects model suggest that there is no significant variation between subgroups defined by language status (monolingual, bilingual or mixed) in terms of the predictive validity of DAs with later word reading outcome measures (Q=0.03, df=2, p=0.98). However, the prediction interval could not be calculated for the bilingual group due to limited number of studies and crossed 0 for mixed language groups but not the monolingual group, suggesting that future relevant studies may be most likely to document positive correlations between DAs and word reading outcomes for monolinguals.

**Table 8.**
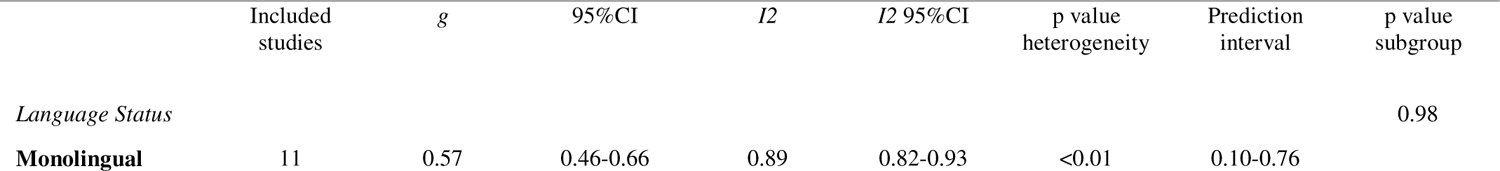
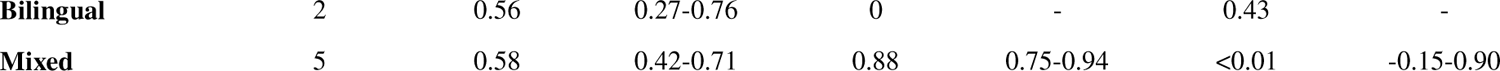
Results of Subgroup Analysis by Language Status for Predictive Validity

|  | Included studies | *g* | 95%CI | *I*<sup>2</sup> | *I*<sup>2</sup> 95%CI | p value heterogeneity | Prediction interval | p value subgroup |
| --- | --- | --- | --- | --- | --- | --- | --- | --- |
| *Language Status* |  |  |  |  |  |  |  | 0.98 |
| **Monolingual** | 11 | 0.57 | 0.46-0.66 | 0.89 | 0.82-0.93 | <0.01 | 0.10-0.76 |  |

|  |  |  |  |  |  |  |  |
| --- | --- | --- | --- | --- | --- | --- | --- |
| **Bilingual** | 2 | 0.56 | 0.27-0.76 | 0 | - | 0.43 | - |
| **Mixed** | 5 | 0.58 | 0.42-0.71 | 0.88 | 0.75-0.94 | <0.01 | -0.15-0.90 |

### Research Question 2B: Do dynamic assessments of word reading skills demonstrate concurrent validity with static assessments of word reading skills, and predictive validity with word reading outcome measures within population groups defined by their reading status (typically developing vs. at-risk)?

#### Concurrent validity –Subgroup analysis reading status

Of the 29 studies that reported a correlation coefficient between a DA and an SA of an equivalent construct 14 included exclusively participants who were typically developing (TD) in their reading abilities 4 included participants who were exclusively at-risk or diagnosed with reading difficulty and 11 included mixed reading status populations in their analyses. Refer to Table 2 for study details. Subgroup analysis by reading status was planned a priori and results are reported in Table 9. The outcomes of this mixed effects model suggest that there is no significant variation between subgroups defined by reading status (typically developing, at-risk or mixed) in terms of the concurrent validity of DAs with their equivalent construct SA counterparts (Q=3.41, df=2, p=0.18). However, the prediction interval crossed 0 for both the at-risk and mixed language groups, but not the typically developing group, suggesting that future relevant studies may be most likely to document positive correlations between DAs and SAs for TD children.

**Table 9.** Results of Subgroup Analysis by Reading Status for Concurrent Validity

|  | Included studies | *g* | 95%CI | *I*<sup>2</sup> | *I*<sup>2</sup> 95%CI | p value heterogeneity | Prediction interval | p value subgroup |
| --- | --- | --- | --- | --- | --- | --- | --- | --- |
| *Reading Status* |  |  |  |  |  |  |  | 0.18 |
| **Typically developing** | 14 | 0.68 | 0.56-0.77 | 0.92 | 0.89-0.95 | <0.01 | 0.09-0.92 |  |
| **At-risk** | 4 | 0.49 | 0.19-0.70 | 0.87 | 0.68-0.95 | <0.01 | -0.78-0.97 |  |
| **Mixed** | 11 | 0.54 | 0.40-0.67 | 0.94 | 0.91-0.96 | <0.01 | -0.13-0.88 |  |

#### Predictive validity – Subgroup analysis reading status

Of the 18 studies that reported a correlation coefficient between a DA and a later word reading outcome measures, 9 studies included exclusively participants who were typically developing, 2 studies included participants who were at-risk or diagnosed with reading difficulty and 7 included populations with mixed reading abilities in their analyses. Please see Table 2 for study details. Subgroup analysis by reading status was planned a priori and results are reported in Table 10. The outcomes of this mixed effects model suggest that there is no significant variation between subgroups defined by reading status (typically developing, at-risk or mixed) in terms of the predictive validity of DAs with later word reading outcome measures (Q=1.35, df=2, p=0.51). Furthermore, the prediction interval did not cross 0 for any subgroup suggesting that future relevant studies are likely to document positive correlations between DAs word reading outcomes for all subgroups regardless of reading status.

**Table 10.**
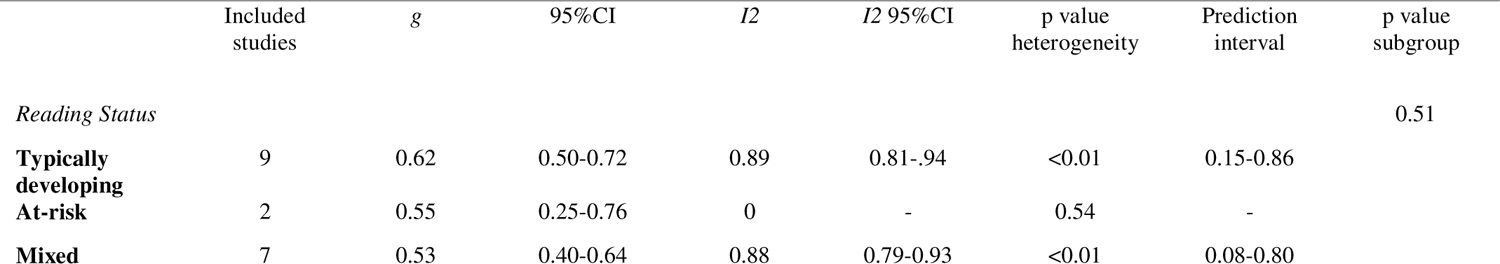
Results of Subgroup Analysis by Reading Status for Predictive Validity

#### Risk of Publication Bias

Two additional analyses were completed to examine potential publication bias for both the concurrent and predictive validity analyses. First, funnel plots were generated. Visual inspection of the funnel plot for the concurrent validity analysis does not suggest asymmetry (Figure 10). Second, Egger’s test was calculated and not found to be significant (Intercept = 1.282, 95%CI= [-1.04 - 3.61], p=.289). Therefore, we conclude that there is no indication of funnel plot asymmetry and minimal risk of publication bias in the analysis for concurrent validity. However, visual inspection of the funnel plot for the predictive validity analysis (Figure 11) suggests potential asymmetry, and Egger’s test was significant for the presence of plot asymmetry (Intercept = 2.4, 95%CI [0.37 - 4.43-, p=.034). Inspection of the plot reveals that there are several small studies with positive effects included in the analysis (e.g., Caffrey, 2006a; Spector, 1992; Horbach et al., 2018), but an absence of smaller studies with negative effects. This suggests that there is a possibility that small studies with negative effects either were not written up, published, or identified in the grey literature search (Lee & Hotopf, 2012).

**Figure 10.**
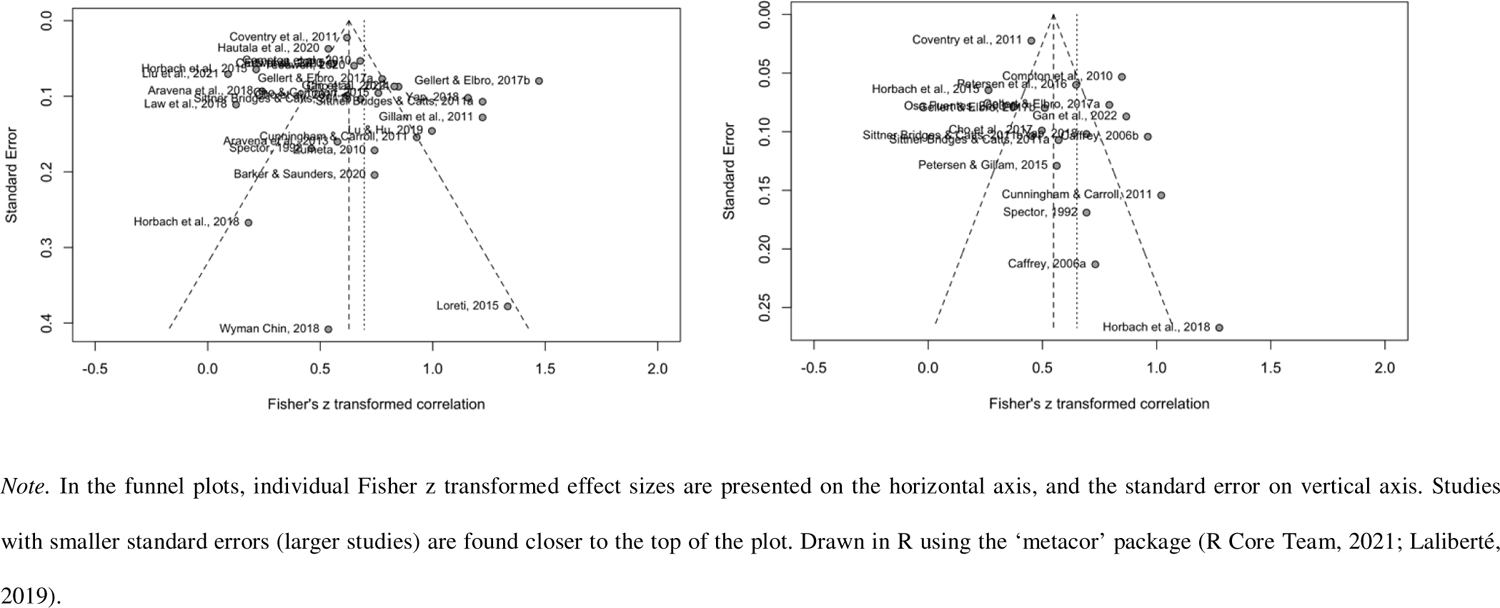
Funnel Plots of Studies Included in the Meta-analysis of the Concurrent Validity Between Dynamic and Static Assessments of Word Reading Skills (Left) And Predictive Validity Between Dynamic Assessments of Word Reading Skills and Word Reading Outcome Measures (Right)

**Figure 11.**
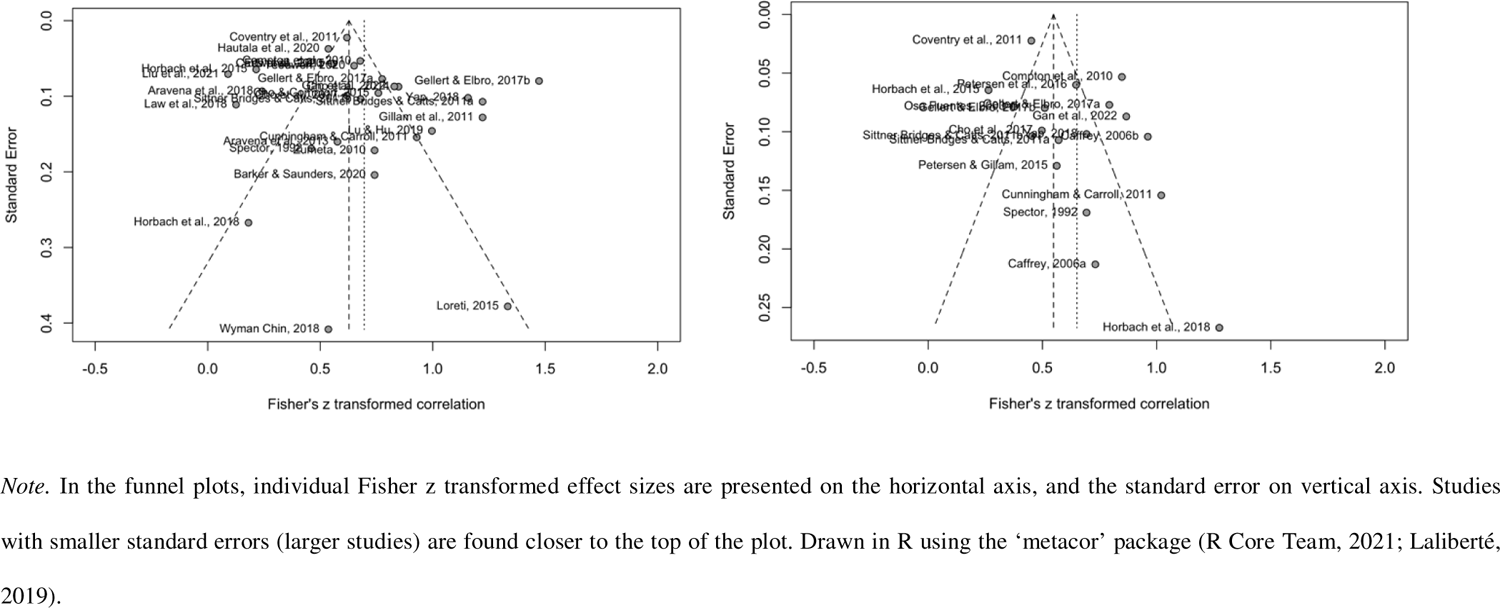
Funnel Plots of Studies Included in the Meta-analysis of the Concurrent Validity Between Dynamic and Static Assessments of Word Reading Skills (Left) And Predictive Validity Between Dynamic Assessments of Word Reading Skills and Word Reading Outcome Measures (Right)

## Discussion

Based on 34 studies from 32 articles, we conducted a systematic review and meta-analysis of the concurrent and predictive validity of dynamic assessments (DA) of word reading skills, including phonological awareness (PA), sound-symbol knowledge (SSK) and decoding. Our meta-analysis examining the *concurrent validity* of DAs with their equivalent static assessment (SA) counterparts suggests that there is a strong correlation between the two types of tests (r=.60). However, subgroup analysis suggests that there are differences in effect sizes for DAs of different literacy skills: DA-PAs demonstrated the strongest correlation and (r=.73) with their SA counterparts and narrowest positive prediction interval, followed by DA-Dec (r=.54) and DA-SSKs (r=.34). There are two possible reasons DAs of PA might demonstrate the strongest concurrent validity with their SA counterparts. First, PA is a well-defined construct with an established hierarchy of skills that is consistent across languages (Anthony & Francis, 2005). Independent of orthography, PA tasks range in complexity of manipulation from simple (e.g., blending) to complex (e.g., manipulation) and in size of unit of speech from larger (e.g., syllables) to smaller (e.g., phonemes). In the included articles, there was a great degree of consistency between the tasks researchers use to evaluate PA dynamically and statically across studies (e.g., a DA of phoneme segmentation compared an SA of phoneme segmentation). Conversely, DAs of SSK and decoding were characterized by a greater degree of variability in task type between DA and SA (e.g., a DA of SSK of learning novel-symbol sound correspondences compared to an SA of letter naming knowledge). Additionally, DAs of PA are likely to correlate more strongly with SAs because they were almost exclusively conducted in-person. This is relevant because most SAs were also conducted in-person and consistency in administration may produce stronger correlations between the two. DAs of SSK, however, were often administered via a computer program and compared to an in-person SA which could have resulted in weaker correlations between the two (e.g., Barker & Saunders, 2020; Horbach et al., 2018).

A second meta-analysis found that that DAs of word reading skills have strong predictive validity with word reading outcome measures (OMs) (r=.57). These findings are consistent with Caffrey et al.,’s 2008 review which documented overall strong effect sizes between DAs and outcome measures across domains. Our findings corroborate Caffrey’s but provide more specific insight into the use of DAs of distinct word reading skills. In the subgroup analysis by DA type, DA-Dec demonstrated the strongest correlation with later word reading outcome measures (r=.59), but this overall weighted effect size was not significantly different from that of the DA-PA with word reading (r=.52). In terms of DA-SSK, only one study was included and could therefore not be compared in subgroup analysis. These findings, which provide suggestive evidence for the validity of DAs of PA and decoding in predicting later word reading outcomes are in line with outcomes of Dixon et al.’s systematic review which found that both DAs of PA and decoding accounted for significant unique variance in later word reading ability (between 1-33% across included studies; Dixon et al., 2022a).

It is unsurprising that DA-Dec showed the strongest correlation in this analysis. A DA that evaluates the ability to learn how to decode words should demonstrate strong predictive validity with later word reading outcomes because these two constructs are similar. It is worth noting that this strong correlation was documented even with variation in the types of DA-Dec and SA-Dec tasks used. For example, some studies used novel symbol nonword decoding task in the DA, and alphabetic real word reading task as an OM and still reported strong correlations between the two (e.g., Gellert & Elbro 2017). These findings complement results from a second systematic review from Dixon et al., which found that DAs targeting constructs that more closely resemble reading ability demonstrated higher classification accuracy in determining reading disorder than those that were more distal (e.g., DAs of decoding better predicted reading status than DAs of morphological awareness).

Significant residual heterogeneity was observed in both the concurrent and predictive validity analyses, even after including the moderator of DA type. Subgroup analyses examining the role of language and reading status were planned a priori to further examine heterogeneity. Regarding *language status* only 9 of 34 studies included bilinguals in their sample, and of those 9, only 4 conducted separate analyses of bilinguals to allow for comparison with monolinguals, while 5 grouped mono and bilinguals together in their analyses. In terms of concurrent validity, subgroup analysis by language status results suggest significant differences in mean effect sizes between groups, with studies conducted with bilinguals demonstrating the strongest correlations between DAs and SAs (r=.78), followed by mixed populations (r=.72) and monolinguals (r=.54). However, these results should be interpreted with caution for two reasons. First there were a limited number of studies included in each of the mixed and bilingual subgroups (n=5 and n=4 respectively) relative to the monolingual group (n=20), a finding which is consistent with previous DA reviews (Dixon et al., 2022a, 2022b). Secondly, the bilingual children in all 4 studies were all typically developing, while the monolingual children varied in terms of their reading status across the 20 studies in this subgroup. These differences could have implications for strength of correlations between DAs and SAs. Participant characteristics cannot be separated from each other in a subgroup analysis but are practically important. A different pattern emerged in the predictive validity analysis by language subgroup, with no significant differences observed between subgroups, suggesting for DAs of word reading demonstrate consistent predictive validity with later reading outcomes across groups regardless of their language status. It should be noted that the nature of bilingualism was often poorly defined in included studies (e.g., sequential vs. simultaneous) and little to no information was provided about the languages spoken, the age of acquisition or proficiency levels. A notable exception is Petersen and Gillam (2015) who evaluated their Spanish/English bilinguals using the Bilingual English Spanish Oral Screener (BESOS; Peña et al., 2009) and reported the number of participants who were English or Spanish dominant versus balanced bilinguals, as well as the average years of expressive language experience in English, Spanish, or bilingual settings.

Subgroup analyses were also conducted to examine whether mean effects sizes differed given participant *reading status* (typically developing vs. at-risk readers). Across the reviewed studies, reading status factored into research questions much more frequently than language status and generally researchers described how at-risk status was defined in their studies. However, only 5 studies conducted separate correlational analyses with at-risk groups or children diagnosed with reading difficulty. An additional 12 studies included mixed groups of participants, and 17 were conducted exclusively with typically developing children. Results of the subgroup analysis for concurrent and predictive validity did not suggest any significant differences in mean effect sizes for groups stratified by their reading status.

The results of the subgroup analyses for predictive validity of DAs stratified by language and reading status differ from findings documented in previous reviews. Specifically, Caffrey et al., (2008) reported that mean effect sizes for DAs and outcome measures (across domains) were strongest for populations with disabilities (r=0.59), followed by typically developing children (r=0.42) and those who were at-risk (r=0.37), while we documented strongest effect sizes for typically developing children (r=0.62), then at-risk groups (r=0.55), then mixed ability groups (r=0.53). This difference may be attributed to the fact that we included only studies examining DAs of word reading skills rather than DAs across cognitive domains or could be a result of separating at-risk and bilingual populations, who were grouped together in the previous review.

### Limitations

Despite a comprehensive database and grey literature search, it is possible that relevant articles were not identified. It is possible that relevant articles were not included because of their language of publication. We included articles published in English, French and Spanish, because these were the languages that we were able to read and extract data from. However, the preprint repository search produced several articles in Portuguese, Korean and Mandarin, that may have also been relevant. Future reviews conducted in the field of bilingual literacy would benefit from purposeful inclusion or cross-cultural and linguistic collaborations with team members who speak and can read other languages so that studies published in non-Western European languages are included. Also, despite the suggestion by Dixon et al., (2022) to consider including PAL in future systematic reviews of DA, our team elected to not include this term in this review primarily because PAL tasks are inherently dynamic in nature.

Finally, we chose correlation coefficients as our measure of effect size to represent the concurrent validity between DAs and SAs and predictive validity between DAs and OMs. Like in the Caffrey et al., review (2008), this choice was made because correlation coefficients were the most observed effect size across studies and allowed for inclusion of a greater number of studies. However, because of this choice, the results can only provide insight into the associations between DAs and SAs or OMs. Though other measures of effect size, like regression coefficients, are better suited to determine causality, the variety in statistical analyses, choice of predictor and outcome variables and study design factors made conducting a regression meta-analysis infeasible.

### Conclusions and Clinical Implications

Despite these limitations, the results of this systematic review and meta-analysis provide suggestive evidence for the concurrent and predictive validity of DAs of phonological awareness (PA) and decoding. DAs of PA and decoding demonstrated strong overall mean effect sizes with SAs of equivalent constructs and with later word reading outcomes. In contrast, there is less evidence to support the validity of DAs of SSK, given that the overall effect size representing the concurrent validity of DAs of SSK with equivalent SAs was only moderate, and there were insufficient studies to conduct a subgroup analysis for the predictive validity of DAs of SSK with later word reading outcomes. Results were mixed regarding the concurrent validity of DAs with SAs across population subgroups, but outcomes indicate that DAs demonstrate strong predictive validity with word reading outcomes across populations, regardless of their language (monolingual/bilingual) or reading status (typically developing/at-risk).

Given these findings, it may be useful for clinicians to incorporate DAs of PA and decoding into their practice. For bilingual children, for whom there are limited word reading screening tools available, DAs may be a suitable alternative to SAs for evaluating and predicting future literacy skills. For monolingual children, clinicians may consider using a DA in addition to an SA. Although this requires additional time, inclusion of a DA in an early literacy assessment battery permits comparison between the outcomes of the two measures. This comparison may help differentiate those who are truly struggling or at-risk for later difficulty from those who lack the skills to perform on the SA, and may be worth the extra time required to administer the DA.

### Future Directions

Future research should examine the validity and use of DAs of word reading skills with well-defined bilingual populations and children who are at-risk for or diagnosed with reading difficulties. DA is often purported to be a less biased method for evaluating the abilities of linguistically diverse children, and as a useful tool for differentiating between those who are truly at-risk for difficulty and those who perform poorly as a result of lack exposure to literacy experiences prior to schooling (e.g., Prath, 2020; Saenz & Huer, 2003). However, this review identified that the majority of studies examining DA to date have been conducted with monolingual children (n=24/34), who are typically-developing (n=17/34).

It will also be critical that these future studies adequately describe the characteristics of their bilingual populations. We noted in this review that while researchers typically described how at-risk status was defined in their articles, even in the rare instances where bilingual children were included in studies, the nature of their bilingualism was poorly defined or not described at all. In the future, researchers should include information about which languages are spoken by bilinguals, report whether these languages were learned simultaneously or sequentially and provide some metric of their proficiency in each language. Without this information, results cannot be meaningfully interpreted for use with any bilingual group.

Beyond language status, it may also be valuable for future studies to examine how other demographic variables contribute to performance on both DAs. For instance, previous studies have indicated that factors like sex, race, and socioeconomic status (SES) contribute to performance on traditional SAs of word reading skills. In the studies included in this review, sex distributions were reported for most studies, but data on participants racial identities and SES backgrounds was limited. It is worth investigating whether DAs have the potential to reduce sex and gender or racial bias in addition to linguistic bias. To achieve this, researchers must consider these intersecting factors in their research design and methodology.

### Author Notes

The authors do not declare any conflicts of interest at the time of publication.

## Data Availability

All data produced in the present study are available upon reasonable request to the authors

## Acknowledgements

This study was supported by a Canada Graduate Scholarship-Master’s grant from the Social Sciences and Humanities Research Council of Canada, at the Rehabilitation Sciences Institute at the University of Toronto and an Ontario Graduate Scholarship from the Ministry of Colleges and Universities awarded to E. Wood, by a University of Toronto Excellence Award, awarded to K. Biggs, and by a Natural Sciences and Engineering Research Council of Canada grant awarded to Dr. M. Molnar.

## Appendix 1.

**Figure 1.**
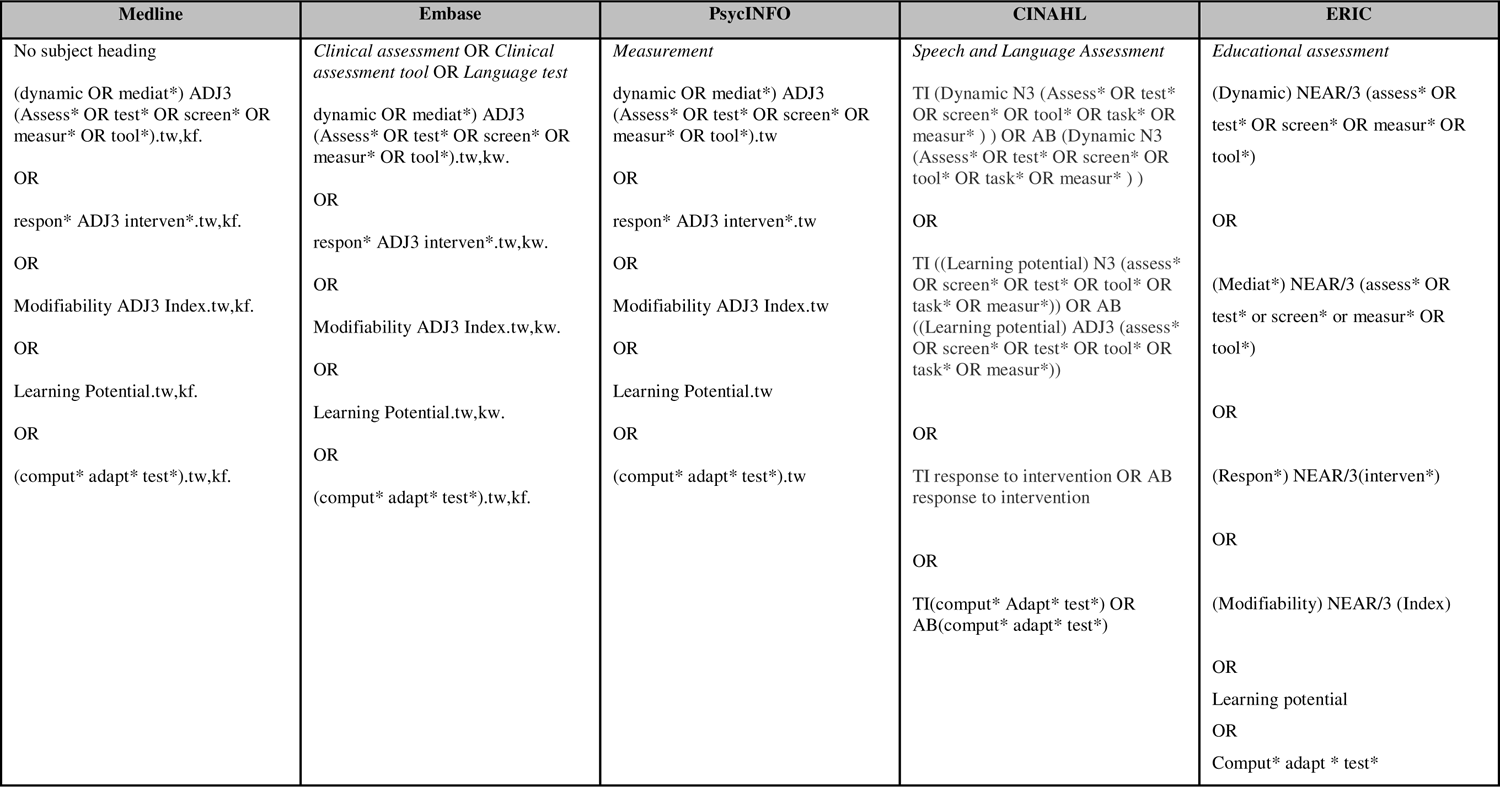
Search terms for concept 1 – Dynamic assessment

**Figure 2.**
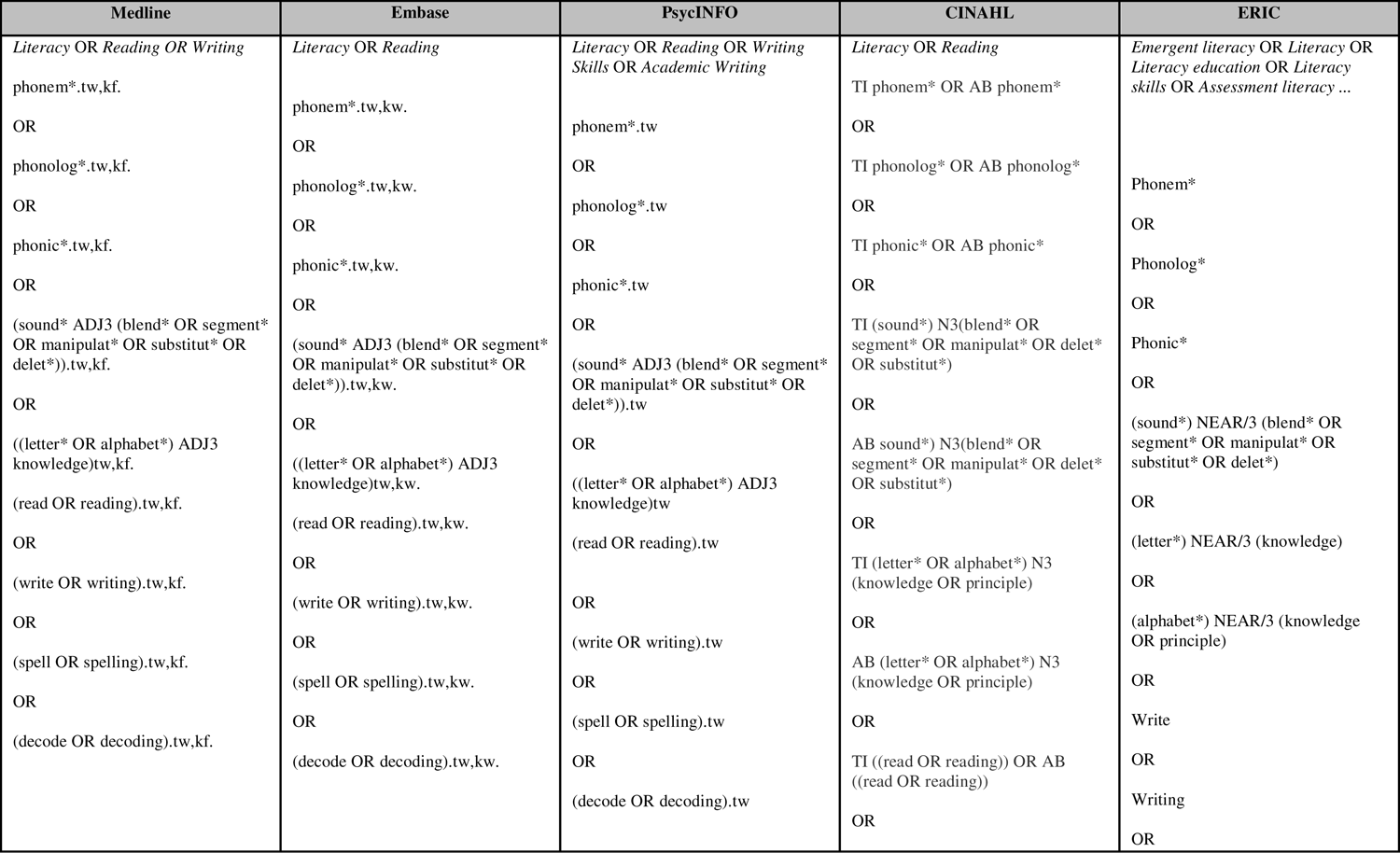
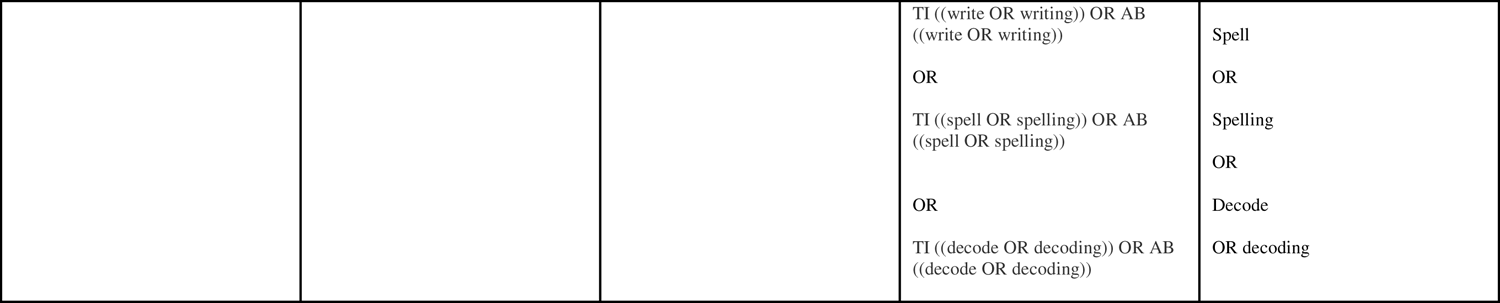
Search terms for concept 2 – Literacy

## Appendix 2.

**Table 3.**
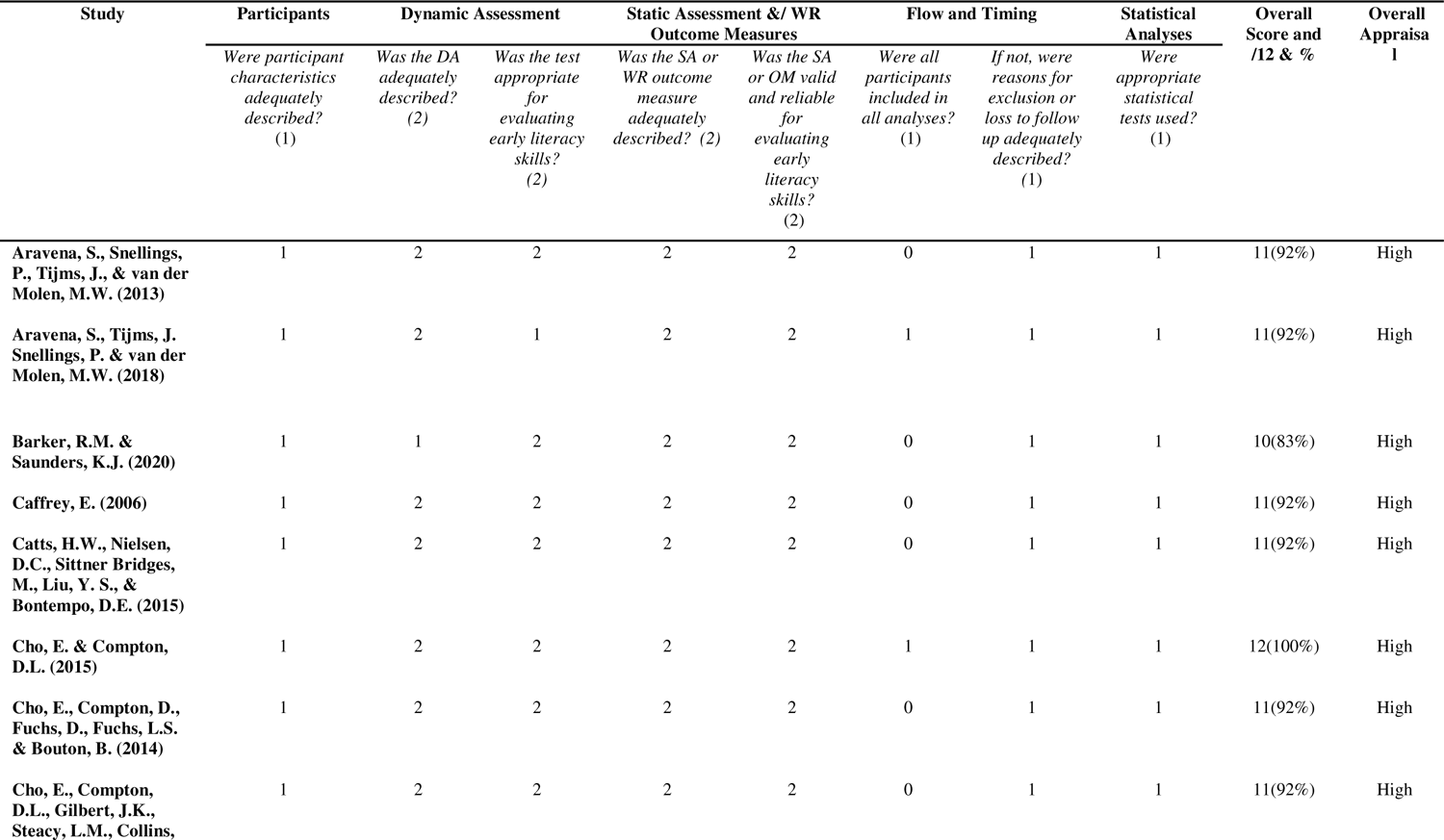
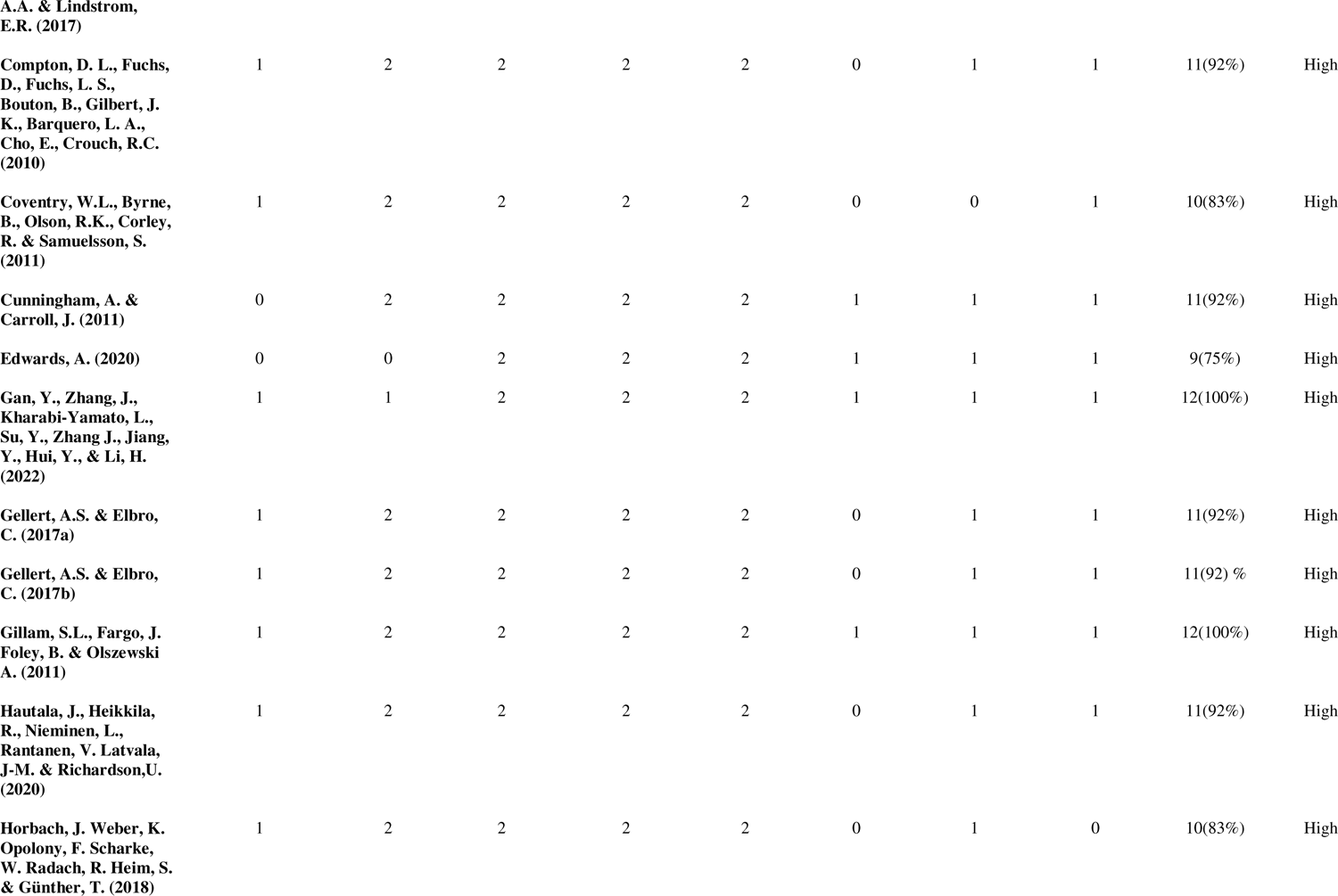
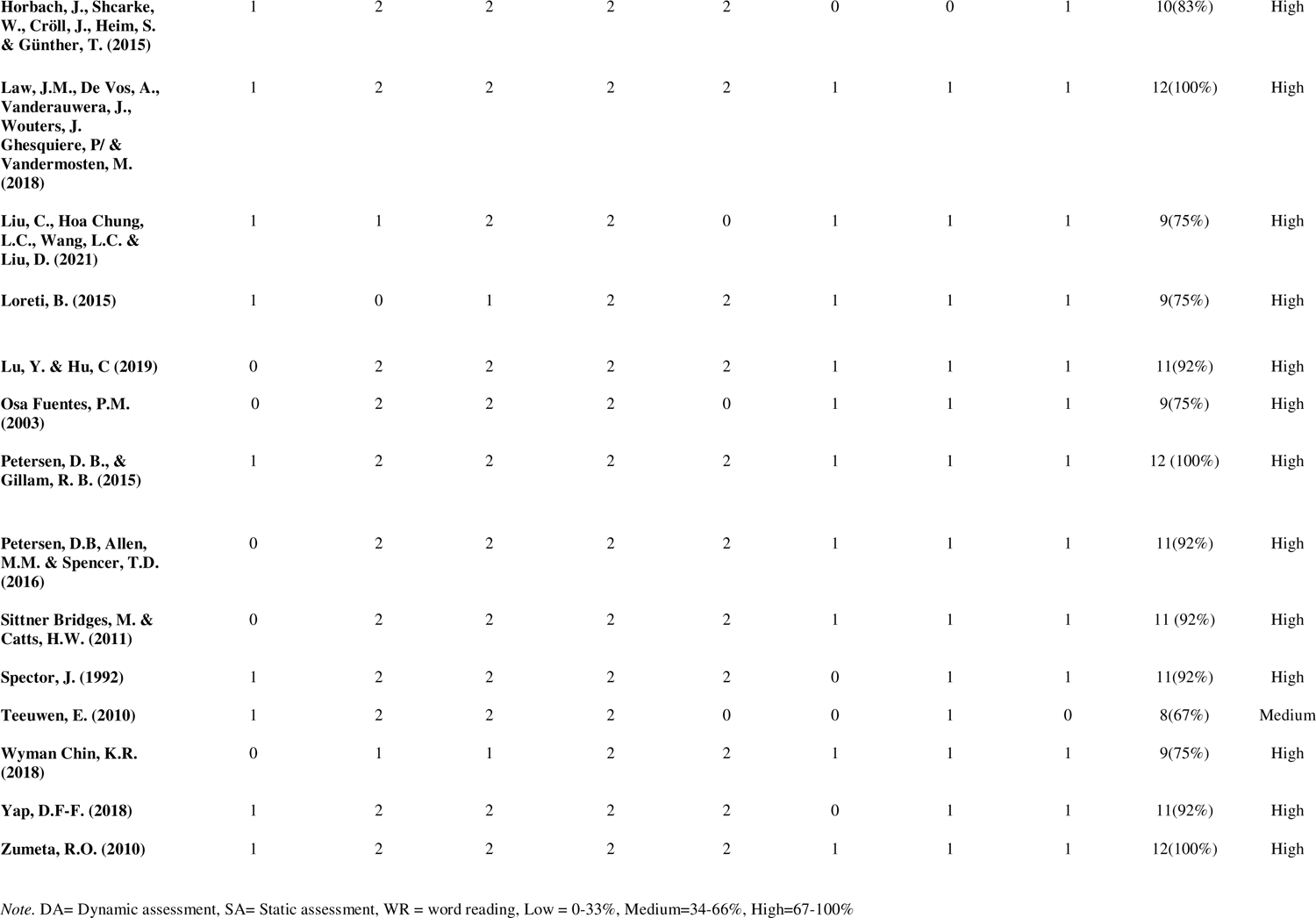
Quality Appraisal of Included Studies

| Study | Participants | Dynamic Assessment |  | Static Assessment &/ WR Outcome Measures |  | Flow and Timing |  | Statistical Analyses | Overall Score and /12 & % | Overall Appraisal |
| --- | --- | --- | --- | --- | --- | --- | --- | --- | --- | --- |
|  | *Were participant characteristics adequately described? (1)* | *Was the DA adequately described? (2)* | *Was the test appropriate for evaluating early literacy skills? (2)* | *Was the SA or WR outcome measure adequately described? (2)* | *Was the SA or OM valid and reliable for evaluating early literacy skills? (2)* | *Were all participants included in all analyses? (1)* | *If not, were reasons for exclusion or loss to follow up adequately described? (1)* | *Were appropriate statistical tests used? (1)* |  |  |
| Aravena, S., Snellings, P., Tijms, J., & van der Molen, M.W. (2013) | 1 | 2 | 2 | 2 | 2 | 0 | 1 | 1 | 11(92%) | High |
| Aravena, S., Tijms, J. Snellings, P. & van der Molen, M.W. (2018) | 1 | 2 | 1 | 2 | 2 | 1 | 1 | 1 | 11(92%) | High |
| Barker, R.M. & Saunders, K.J. (2020) | 1 | 1 | 2 | 2 | 2 | 0 | 1 | 1 | 10(83%) | High |
| Caffrey, E. (2006) | 1 | 2 | 2 | 2 | 2 | 0 | 1 | 1 | 11(92%) | High |
| Catts, H.W., Nielsen, D.C., Sittner Bridges, M., Liu, Y. S., & Bontempo, D.E. (2015) | 1 | 2 | 2 | 2 | 2 | 0 | 1 | 1 | 11(92%) | High |
| Cho, E. & Compton, D.L. (2015) | 1 | 2 | 2 | 2 | 2 | 1 | 1 | 1 | 12(100%) | High |
| Cho, E., Compton, D., Fuchs, D., Fuchs, L.S. & Bouton, B. (2014) | 1 | 2 | 2 | 2 | 2 | 0 | 1 | 1 | 11(92%) | High |
| Cho, E., Compton, D.L., Gilbert, J.K., Steacy, L.M., Collins, | 1 | 2 | 2 | 2 | 2 | 0 | 1 | 1 | 11(92%) | High |
| **A.A. & Lindstrom, E.R. (2017)** |  |  |  |  |  |  |  |  |  |  |
| **Compton, D. L., Fuchs, D., Fuchs, L. S., Bouton, B., Gilbert, J. K., Barquero, L. A., Cho, E., Crouch, R.C. (2010)** | 1 | 2 | 2 | 2 | 2 | 0 | 1 | 1 | 11(92%) | High |
| **Coventry, W.L., Byrne, B., Olson, R.K., Corley, R. & Samuelsson, S. (2011)** | 1 | 2 | 2 | 2 | 2 | 0 | 0 | 1 | 10(83%) | High |
| **Cunningham, A. & Carroll, J. (2011)** | 0 | 2 | 2 | 2 | 2 | 1 | 1 | 1 | 11(92%) | High |
| **Edwards, A. (2020)** | 0 | 0 | 2 | 2 | 2 | 1 | 1 | 1 | 9(75%) | High |
| **Gan, Y., Zhang, J., Kharabi-Yamato, L., Su, Y., Zhang J., Jiang, Y., Hui, Y., & Li, H. (2022)** | 1 | 1 | 2 | 2 | 2 | 1 | 1 | 1 | 12(100%) | High |
| **Gellert, A.S. & Elbro, C. (2017a)** | 1 | 2 | 2 | 2 | 2 | 0 | 1 | 1 | 11(92%) | High |
| **Gellert, A.S. & Elbro, C. (2017b)** | 1 | 2 | 2 | 2 | 2 | 0 | 1 | 1 | 11(92) % | High |
| **Gillam, S.L., Fargo, J. Foley, B. & Olszewski A. (2011)** | 1 | 2 | 2 | 2 | 2 | 1 | 1 | 1 | 12(100%) | High |
| **Hautala, J., Heikkila, R., Nieminen, L., Rantanen, V. Latvala, J-M. & Richardson,U. (2020)** | 1 | 2 | 2 | 2 | 2 | 0 | 1 | 1 | 11(92%) | High |
| **Horbach, J. Weber, K. Opolony, F. Scharke, W. Radach, R. Heim, S. & Günther, T. (2018)** | 1 | 2 | 2 | 2 | 2 | 0 | 1 | 0 | 10(83%) | High |
| **Horbach, J., Shcarke, W., Cröll, J., Heim, S. & Günther, T. (2015)** | 1 | 2 | 2 | 2 | 2 | 0 | 0 | 1 | 10(83%) | High |
| **Law, J.M., De Vos, A., Vanderauwera, J., Wouters, J. Ghesquiere, P/ & Vandermosten, M. (2018)** | 1 | 2 | 2 | 2 | 2 | 1 | 1 | 1 | 12(100%) | High |
| **Liu, C., Hoa Chung, L.C., Wang, L.C. & Liu, D. (2021)** | 1 | 1 | 2 | 2 | 0 | 1 | 1 | 1 | 9(75%) | High |
| **Loreti, B. (2015)** | 1 | 0 | 1 | 2 | 2 | 1 | 1 | 1 | 9(75%) | High |
| **Lu, Y. & Hu, C (2019)** | 0 | 2 | 2 | 2 | 2 | 1 | 1 | 1 | 11(92%) | High |
| **Osa Fuentes, P.M. (2003)** | 0 | 2 | 2 | 2 | 0 | 1 | 1 | 1 | 9(75%) | High |
| **Petersen, D. B., & Gillam, R. B. (2015)** | 1 | 2 | 2 | 2 | 2 | 1 | 1 | 1 | 12 (100%) | High |
| **Petersen, D.B, Allen, M.M. & Spencer, T.D. (2016)** | 0 | 2 | 2 | 2 | 2 | 1 | 1 | 1 | 11(92%) | High |
| **Sittner Bridges, M. & Catts, H.W. (2011)** | 0 | 2 | 2 | 2 | 2 | 1 | 1 | 1 | 11 (92%) | High |
| **Spector, J. (1992)** | 1 | 2 | 2 | 2 | 2 | 0 | 1 | 1 | 11(92%) | High |
| **Teeuwen, E. (2010)** | 1 | 2 | 2 | 2 | 0 | 0 | 1 | 0 | 8(67%) | Medium |
| **Wyman Chin, K.R. (2018)** | 0 | 1 | 1 | 2 | 2 | 1 | 1 | 1 | 9(75%) | High |
| **Yap, D.F-F. (2018)** | 1 | 2 | 2 | 2 | 2 | 0 | 1 | 1 | 11(92%) | High |
| **Zumeta, R.O. (2010)** | 1 | 2 | 2 | 2 | 2 | 1 | 1 | 1 | 12(100%) | High |

*Note.* DA= Dynamic assessment, SA= Static assessment, WR = word reading, Low = 0-33%, Medium=34-66%, High=67-100%

## Appendix 3.

**Figure 12.**
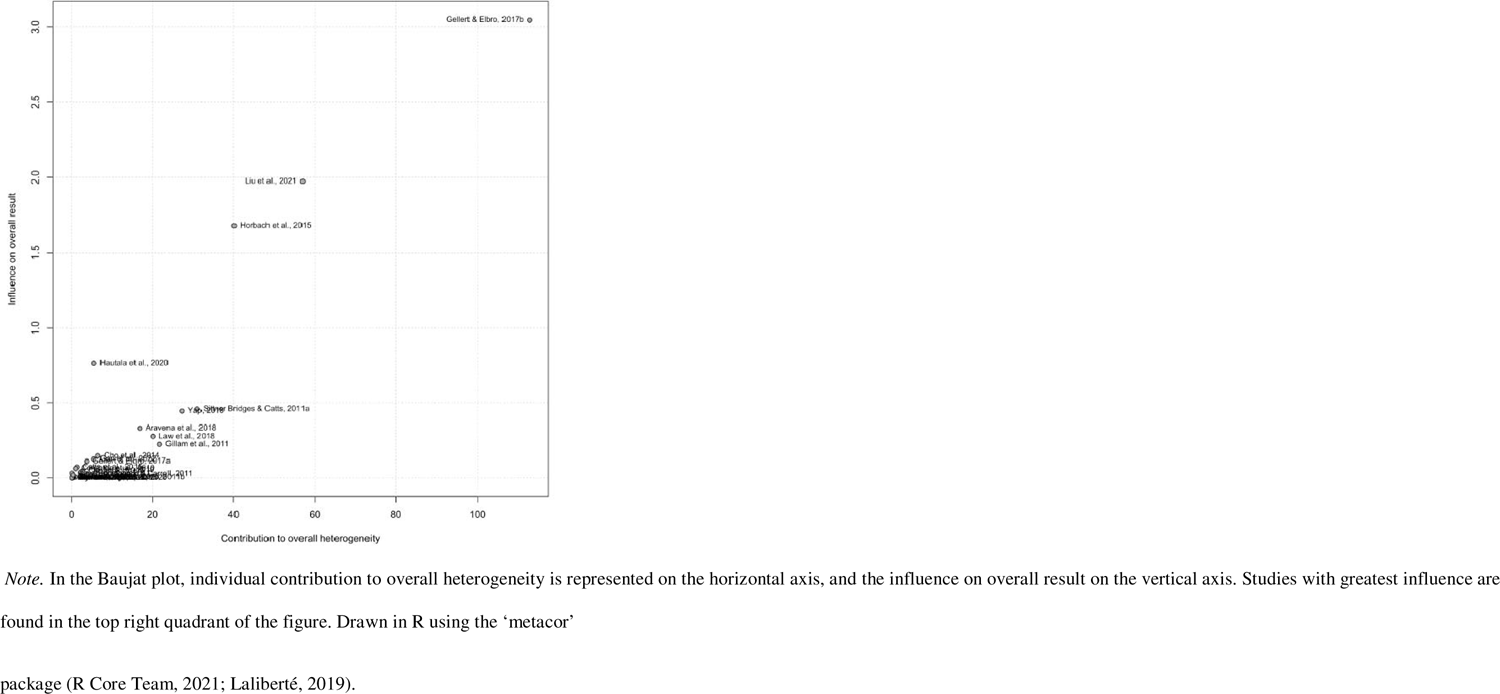
Baujat plot for studies included in the meta-analysis of the concurrent validity of dynamic and static assessments of early literacy skills

**Figure 13.**
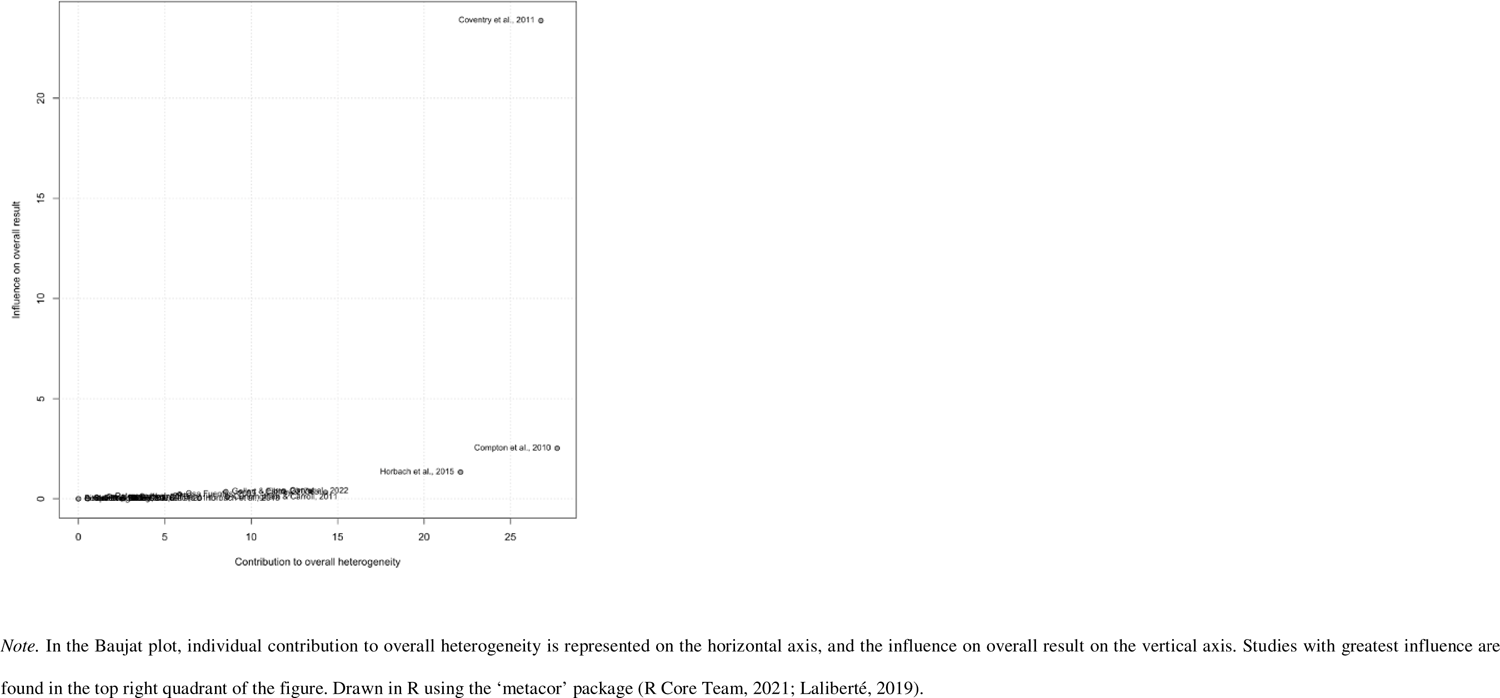
Baujat plot for studies included in the meta-analysis of the predictive validity of dynamic assessments of early literacy skills with word reading outcome measures

## Footnotes

1 Coders also reported whether the DA was administered in the traditional “in-person” format, or via computer, which type of DA was used, the Train/Test or Graduated Prompt format, and whether the DA used real words or nonwords, familiar letters/characters of novel symbols. Results of the comparisons between in-person and computerized DA, train/test vs. Graduated prompts DA, word/nonword and familiar/novel symbol will be reported in a separate forthcoming study regarding the relationship between the different characteristics of DAs of word reading skills and word reading measures.

