## Appendices for "Concurrent and predictive validity of dynamic assessments of word reading in young children: A systematic review and meta-analysis" (from another version of this preprint: version 1, 2022-09-20)

### Appendix 1.

**Figure 8.**

*Search terms for concept 1 – Dynamic assessment*

| Medline | Embase | PsycINFO | ERIC |
|---|---|---|---|
| No subject heading | *Clinical assessment* OR *Clinical assessment tool* OR *Language test* | *Measurement* | *Educational assessment* |
| (dynamic OR mediat*) ADJ3 (Assess* OR test* OR screen* OR measur* OR tool*).tw,kf. | dynamic OR mediat*) ADJ3 (Assess* OR test* OR screen* OR measur* OR tool*).tw,kw. | dynamic OR mediat*) ADJ3 (Assess* OR test* OR screen* OR measur* OR tool*).tw | Dynamic) NEAR/3 (assess* OR test* OR screen* OR measur* OR tool*) |
| OR | OR | OR | OR |
| respon* ADJ3 interven*.tw,kf. | respon* ADJ3 interven*.tw,kw. | respon* ADJ3 interven*.tw | (Mediat*) NEAR/3 (assess* OR test* or screen* or measur* OR tool*) |
| OR | OR | OR | OR |
| Modifiability ADJ3 Index.tw,kf. | Modifiability ADJ3 Index.tw,kw. | Modifiability ADJ3 Index.tw | (Respon*) NEAR/3(interven*) |
| OR | OR | OR | OR |
| Learning Potential.tw,kf. | Learning Potential.tw,kw. | Learning Potential.tw | |

| | | | |
|---|---|---|---|
| | | | (Modifiability) NEAR/3 (Index)<br><br>OR<br><br>Learning potential |

*Note.* An additional search term (comput* adapt* test*) was added in a second round of database searching as an additional synonym for dynamic assessment upon identifying several articles that used this term.

**Figure 9.**

*Search terms for concept 2 – Literacy*

| Medline | Embase | PsycINFO | ERIC |
|---|---|---|---|
| *Literacy* OR *Reading OR Writing*<br><br>phonem*.tw,kf.<br><br>OR<br><br>phonolog*.tw,kf.<br><br>OR | *Literacy* OR *Reading*<br><br>phonem*.tw,kw.<br><br>OR<br><br>phonolog*.tw,kw.<br><br>OR | *Literacy* OR *Reading* OR *Writing Skills* OR *Academic Writing*<br><br>phonem*.tw<br><br>OR<br><br>phonolog*.tw<br><br>OR | *Emergent literacy* OR *Literacy* OR *Literacy education* OR *Literacy skills* OR *Assessment literacy ...*<br><br>Phonem*<br><br>OR<br><br>Phonolog*<br><br>OR |

| | | | |
|---|---|---|---|
| phonic*.tw,kf.<br><br>OR<br><br>(sound* ADJ3 (blend* OR segment* OR manipulat* OR substitut* OR delet*)).tw,kf.<br><br>OR<br><br>((letter* OR alphabet*) ADJ3 knowledge)tw,kf.<br><br>(read OR reading).tw,kf.<br><br>OR<br><br>(write OR writing).tw,kf.<br><br>OR<br><br>(spell OR spelling).tw,kf. | phonic*.tw,kw.<br><br>OR<br><br>(sound* ADJ3 (blend* OR segment* OR manipulat* OR substitut* OR delet*)).tw,kw.<br><br>OR<br><br>((letter* OR alphabet*) ADJ3 knowledge)tw,kw.<br><br>(read OR reading).tw,kw.<br><br>OR<br><br>(write OR writing).tw,kw.<br><br>OR<br><br>(spell OR spelling).tw,kw. | phonic*.tw<br><br>OR<br><br>(sound* ADJ3 (blend* OR segment* OR manipulat* OR substitut* OR delet*)).tw<br><br>OR<br><br>((letter* OR alphabet*) ADJ3 knowledge)tw<br><br>(read OR reading).tw<br><br>OR<br><br>(write OR writing).tw<br><br>OR | Phonic*<br><br>OR<br><br>(sound*) NEAR/3 (blend* OR segment* OR manipulat* OR substitut* OR delet*)<br><br>OR<br><br>(letter*) NEAR/3 (knowledge)<br><br>OR<br><br>(alphabet*) NEAR/3 (knowledge OR principle)<br><br>OR<br><br>Write<br><br>OR |

| | | | |
|---|---|---|---|
| OR<br><br>(decode OR decoding).tw,kf. | OR<br><br>(decode OR decoding).tw,kw. | (spell OR spelling).tw<br><br>OR<br><br>(decode OR decoding).tw | Writing<br><br>OR<br><br>Spell<br><br>OR<br><br>Spelling<br><br>OR<br><br>Decode<br><br>OR decoding |

### Appendix 2.

**Table 1.**

***General Characteristics of Each Study***

| **Study** | Country | N | Mean age a start (Years; months) | Grade at start | Gender | Language Status & Language Spoken | Reading Status | Timing |
|---|---|---|---|---|---|---|---|---|
| **Aravena, S., Snellings, P., Tijms, J., & van der Molen, M. W. (2013)** | The Netherlands | 64 TD<br>62 Dx.<br>42 Dx. * | 9;9 TD<br>9;9 Dx. | Not stated | 48.4% males TD<br>56.4% males Dx. | ML Danish | TD & Dx | Cross-sectional |
| **Aravena, S., Tijms, J. Snellings, P. & van der Molen, M.W. (2018)** | The Netherlands | 46 TD<br>72 Dx.<br>71 Dx. * | 9;3 TD<br>9;2 Dx. | Not stated | 47.8% males TD<br>58.2% males Dx. | ML Danish | TD & Dx | Cross-sectional |
| **Barker, R.M. & Saunders, K.J. (2020)** | United States | 27 | 4;11 | Preschool | 51.8% males | ML English | TD | Cross-sectional |

| | | | | | | | | |
|---|---|---|---|---|---|---|---|---|
| **Caffrey, E. (2006)** | United States | 120 K<br>95 G1 | Not stated | K<br>G1 | 52% males K<br>57% males G1 | ML English | TD | Longitudinal |
| **Catts, H.W., Nielsen, D.C., Sittner Bridges, M., Liu, Y. S., & Bontempo, D.E. (2015)** | United States | 103 TD<br>263 AR<br>313 total | Not stated | K | 51.4% males TD<br>57% males AR | ML English | TD & AR | Cross-sectional |
| **Cho, E., Compton, D., Fuchs, D., Fuchs, L.S. & Bouton, B. (2014)** | United States | 134 | Not stated | G1 | 50% males | ML English | AR | Cross-Sectional |
| **Cho, E. & Compton, D.L. (2015)** | United States | 112 | 6;8 | G1 | 57.1% male | ML English | TD | Cross-sectional |
| **Cho, E., Compton, D.L., Gilbert, J.K., Steacy, L.M., Collins, A.A. & Lindstrom, E.R. (2017)** | United States | 54 TD<br>51 AR<br>105 total | 6;7 TD<br>6;10 AR<br>6;8 | G1 | 55% males | ML English | TD & AR | Cross-Sectional<br>Longitudinal |
| **Clayton, F. (2018)** | England | 97 | 9;2 | Year 4<br>Year 5 | 50.5% males | ML English | TD | Cross-Sectional |
| **Compton, D. L., Fuchs, D., Fuchs, L. S., Bouton, B., Gilbert, J. K.,** | United States | 140 TD<br>214 AR | Not stated | 1 | 53.45% males total | ML English | TD & AR | Cross-Sectional<br>Longitudinal |

| | | | | | | | | |
|---|---|---|---|---|---|---|---|---|
| **Barquero, L. A., Cho, E., Crouch, R.C. (2010)** | | 354 total | | | | | | |
| **Coventry, W.L., Byrne, B., Olson, R.K., Corley, R. & Samuelsson, S. (2011)** | Australia<br>United States<br>Norway<br>Sweden | 1988 | 4;11 | Preschool | 48% males MZ<br>53% males DZ | ML English<br>ML Norwegian<br>ML Swedish<br>BL not specified | TD | Cross-Sectional<br>Longitudinal |
| **Cunningham, A. & Carroll, J. (2011)** | England | 45 | 5;0 K<br>5.2 young G1<br>5;11 old G1 | K<br>G1 | Not stated | ML English | TD | Cross-sectional<br>Longitudinal |
| **Edwards, A. (2020)** | United States | 312 | Not stated | G1 | Not stated | ML English | TD | Cross-Sectional<br>Longitudinal |
| **Gellert, A.S. & Elbro, C. (2017a)** | Denmark | 89 TD<br>82 AR<br>171 total | 6;11 | K | Not stated | ML Danish-110<br>BL not specified 61 | TD & AR | Cross-Sectional<br>Longitudinal |
| **Gellert, A.S. & Elbro, C. (2017b)** | Denmark | 84 TD<br>76 AR | 6;4 | K | 48% males | ML Danish - 101 | TD & AR | Cross-Sectional<br>Longitudinal |

| | | | | | | | | |
|---|---|---|---|---|---|---|---|---|
| | | 160 total | | | | BL not specified 59 | | |
| **Gillam, S.L., Fargo, J. Foley, B. & Olszewski, A. (2011)** | United States | 64 | 6;5 young<br>8;0 old | G1<br>G2<br>G3 | 47% males | ML English | TD | Cross-Sectional |
| **Hautala, J., Heikkila, R., Nieminen, L., Rantanen, V. Latvala, J-M. & Richardson, U. (2020)** | Finland | 723 | Not stated | G1<br>G2<br>G3<br>G4 | 44.5% males | ML Finnish | TD & AR | Cross-Sectional |
| **Horbach, J., Shcarke, W., Cröll, J., Heim, S. & Günther, T. (2015)** | Germany | 243 | 6;2 | K | 56.0% male | ML German | TD & AR | Cross-Sectional<br>Longitudinal |
| **Horbach, J. Weber, K. Opolony, F. Scharke, W. Radach, R. Heim, S. & Günther, T. (2018)** | Germany | 17 | 5;0 | Preschool | 41% males | ML German-6<br>BL not specified 11 | TD | Cross-Sectional<br>Longitudinal |
| **Law, J.M., De Vos, A., Vanderauwera, J., Wouters, J. Ghesquiere, P/ & Vandermosten, M. (2018)** | Belgium | 64 TD<br>20 Dx.<br>84 total | 8;3 | G3 | Not stated | ML Dutch | TD & Dx. | Cross-Sectional |

| | | | | | | | | |
|---|---|---|---|---|---|---|---|---|
| **Liu, C., Hoa Chung, K.K., Wang, L.C. & Liu, D. (2021)** | China | 203 | 5;0 | K Year 2 | 51.23% males | ML Mandarin | TD | Cross-Sectional |
| **Lu, Y. & Hu, C (2019)** | Taiwan | 50 | Not stated | G4 | Not stated | BL Mandarin & English | TD | Cross-Sectional |
| **Osa Fuentes, P.M. (2003)** | Spain | 164 | 5;6 | Infantil (Preschool or K equivalent) | 45.7% males | ML Spanish | TD | Longitudinal |
| **Petersen, D. B., & Gillam, R. B. (2015)** | United States | 63 | 5;4 | K | 53.9% males | BL Spanish & English | AR | Longitudinal |
| **Petersen, D.B, Allen, M.M. & Spencer, T.D. (2016)** | United States | 280 | Not stated | K | Not stated | ML English<br>BL Spanish & English | TD & AR | Longitudinal |
| **Sittner Bridges, M. & Catts, H.W. (2011) (Sample 1)** | United States | 90 | Not stated | K | Not stated | ML English | TD | Cross-Sectional<br>Longitudinal |
| **Sittner Bridges, M. & Catts, H.W. (2011) (Sample 2)** | United States | 96 | Not stated | K | Not stated | ML English | TD & AR | Cross-Sectional<br>Longitudinal |
| **Spector, J. (1992)** | United States | 38 | 5;11 | K | Not stated | ML English | AR | Cross-Sectional |

| | | | | | | | | |
|---|---|---|---|---|---|---|---|---|
| | | | | | | | | Longitudinal |
| **Teeuwen, E. (2020)** | The Netherlands | 284 | 5;5 | K | 52% males | ML Dutch | TD & AR | Cross-sectional |
| **Yap, D.F-F. (2018)** | Singapore | 99 | 5;3 | K | 43.4% males | BL English & Chinese (75%) -Malay (12%) -Other (13%) | TD | Cross-Sectional Longitudinal |
| **Zumeta, R.O. (2010)** | United States | 37 | 6;1 | K | 51.4% male | ML English BL not specified | TD & AR | Cross-sectional |

*Note*. TD= typically developing, AR= at-risk, Dx. = diagnosed with difficulty, G=grade, K = kindergarten ML = monolingual, BL=bilingual, N= number of participants per study

**Table 2.**

***Characteristics of Assessments***

| **Study** | Skill(s) Evaluated with DA | Feedback in DA | Prompting in DA | Scoring of DA | Skill(s) Evaluated with SA | SA Measure(s) Used | Outcome Skill(s) Evaluated | Outcome Measure(s) Used |
|---|---|---|---|---|---|---|---|---|
| **Aravena, S., Snellings, P., Tijms, J., & van der Molen, M.W. (2013)** | SSK<br>-learn novel symbol-sound corresponden ce<br>[explicit/ implicit]<br>Decoding<br>-read real words comprised of novel symbols | Implicit<br>-correct responses lead to game success<br>[implicit condition] | N/A | SSK<br>-# of correct responses<br>Decoding<br>-# of words read correctly | SSK<br>-letter name knowledge<br>Decoding<br>-word reading accuracy | Researcher developed tools<br>One-Minute Test | N/A | N/A |
| **Aravena, S., Tijms, J. Snellings, P. & van der Molen, M.W. (2018)** | SSK<br>-learn novel symbol-sound corresponden ce [implicit]<br>Decoding | Implicit<br>-correct responses lead to game success | N/A | SSK<br>-# of correct responses<br>[accuracy]<br>-median of latencies for correct | PA<br>-phoneme deletion<br>Decoding | 3DM | N/A | N/A |

| | | | | | | | | |
|---|---|---|---|---|---|---|---|---|
| | -read real words comprised of novel symbols | | | response [speed]<br><br>Decoding –% of words read correctly [accuracy<br><br>-#of words read correctly in 30s<br><br>[speed] | -word reading accuracy<br><br>-word reading speed | | | |
| **Barker, R.M. & Saunders, K.J. (2020)** | SSK<br><br>-learn novel symbol-syllable corresponden ce<br><br>[explicit] | Explicit verbal | Teach trials<br><br>-visual support for audio-visual matching | Graded<br><br>-maximum 3 points per item (6 items per subtest)<br><br>-maximum 3 points per subtest (item scores averaged across 4 subtests)<br><br>-maximum 12 points total (sum of 4 subtests)<br><br>-higher score = better | PA<br><br>-phoneme deletion<br><br>-phoneme blending<br><br>-phoneme matching<br><br>SSK<br><br>-letter sound knowledge<br><br>Decoding<br><br>-untimed word reading | CTOPP-2<br><br>-elision<br><br>-phoneme blending<br><br>-sound matching<br><br>Curriculum based Measurement for Early Literacy<br><br>-letter sound knowledge<br><br>WRMT-III | N/A | N/A |

| | | | | | | | | |
|---|---|---|---|---|---|---|---|---|
| | | | | | -untimed nonword reading | -word identification<br>-word attack | | |
| **Caffrey, E. (2006)** | Decoding<br>- read nonwords<br>(CVC, CVCe, CVCCing) | Explicit verbal | Graded<br>-5 levels of increasingly explicit prompts | Graded<br>-maximum 5 points per level<br>(3 levels)<br>-higher score = worse | SSK<br>- letter & grapheme sound knowledge<br>Decoding<br>-untimed word reading | Curriculum based measures<br>-letter sound knowledge<br>WRAT<br>-letter name knowledge<br>-word reading subtest<br>WRMT-R<br>-word attack | WR<br>NWR<br>WW | WRAT<br>-word reading subtest<br>WRMT-R<br>-word identification<br>-word attack<br>WIAT<br>-spelling subtest |
| **Catts, H.W., Nielsen, D.C., Sittner Bridges, M., Liu, Y. S., & Bontempo, D.E. (2015)** | PA<br>-syllable/ phoneme deletion real words | Explicit verbal | Graded<br>-3 levels of increasingly explicit prompts | Graded<br>-maximum 4 points per item -higher sore = better | PA<br>-phoneme<br>Matching<br>-phoneme identification fluency | CTOPP<br>-sound matching<br>DIBELS<br>-initial sound fluency | WR<br>NWR<br>PR | WRMT-R/NU<br>-word identification<br>-word attack<br>TOWRE-2 |

| | | | | | | | | |
|---|---|---|---|---|---|---|---|---|
| | | | | | SSK<br>-letter naming fluency | -letter naming fluency | | -sight word efficiency<br>-phonemic decoding efficiency<br><br>FAIR<br>-oral reading fluency |
| **Cho, E., Compton, D., Fuchs, D., Fuchs, L.S. & Bouton, B. (2014)** | Decoding<br>-read nonwords (CVC, CVCe and CVCCing) | Explicit Verbal | Graded<br>-5 levels of increasingly explicit prompts | Graded<br>-maximum 5 points per level<br>(3 levels)<br>-higher score = worse | PA<br>-phoneme matching<br><br>Decoding<br>-untimed nonword reading<br>-timed nonword reading | CTOPP<br>-sound matching<br><br>WRMT-R/NU<br>-word attack<br><br>TOWRE<br>-phonemic decoding efficiency | N/A | N/A |
| **Cho, E. & Compton, D.L. (2015)** | SSK<br>-learn novel symbol-sound | SSK | Graded<br>-varying levels of increasingly | Graded<br>-total number of correct items across | PA<br>-phoneme deletion | CTOPP<br>-elision | N/A | N/A |

| | | | | | | | | |
|---|---|---|---|---|---|---|---|---|
| | correspondence [explicit]<br><br>Decoding<br>-read nonwords comprised of novel symbols<br>(CVC)<br>-read nonwords comprised of novel symbols and infer silent e rule (CVCe) | -Explicit verbal and visual<br><br>Decoding<br>-CVC- Explicit verbal<br>-CVCe- Explicit verbal and visual | explicit prompts<br><br>SSK<br>-9 levels<br><br>Decoding<br>-CVC – 4 levels<br>-CVCe –5 levels | mastery tests (14)<br>-higher score = better | Decoding<br>-untimed word reading<br>-timed word reading<br>-untimed novel symbol word reading | WRMT – R/NU: -word attack<br><br>TOWRE<br>-phonemic decoding efficiency<br><br>Research developed tools<br>-read 12 English words made up of novel symbols | | |
| **Cho, E., Compton, D.L., Gilbert, J.K., Steacy, L.M., Collins, A.A. & Lindstrom, E.R. (2017)** | SSK<br>-learn novel symbol-sound correspondence [explicit]<br><br>Decoding<br>-read nonwords comprised of | SSK<br>-Explicit verbal and visual<br><br>Decoding<br>-CVC- Explicit verbal<br>-CVCe- Explicit | Graded<br>-varying levels of increasingly explicit prompts<br><br>SSK<br>-9 levels<br>Decoding | Graded<br>-total number of correct items across mastery tests (14)<br>-higher score = better | PA<br>-phoneme deletion<br><br>Decoding<br>-untimed word reading<br>-untimed nonword reading | CTOPP<br>-elision<br><br>WRMT<br>-word attack<br>-word identification<br><br>TOWRE | WR<br>NWR | WRMT<br>-word attack<br>-word identification<br><br>TOWRE<br>-sight word efficiency |

| | | | | | | | | |
|---|---|---|---|---|---|---|---|---|
| | novel symbols<br>(CVC)<br>-read nonwords comprised of novel symbols and infer silent e rule (CVCe) | verbal and visual | -CVC – 4 levels<br>-CVCe –5 levels | | -timed word reading<br>-timed nonword reading | -sight word efficiency<br>-phonemic decoding efficiency | | -phonemic decoding efficiency |
| **Clayton, F. (2018)** | SSK<br>-learn novel symbol-sound and symbol-syllable correspondence | Explicit verbal | N/A | # of correct items on test blocks | Decoding<br>-untimed word reading<br>-timed word reading<br>-timed nonword reading | SWRT<br>-single word reading accuracy<br>TOWRE<br>-sight word reading efficiency<br>-phonemic decoding efficiency | N/A | N/A |

| | | | | | | | | |
|---|---|---|---|---|---|---|---|---|
| **Compton, D. L., Fuchs, D., Fuchs, L. S., Bouton, B., Gilbert, J. K., Barquero, L. A., Cho, E., Crouch, R.C. (2010)** | Decoding<br>-read nonwords<br>(CVC, CVCe, CVCCing) | Explicit Verbal | Graded<br>- 5 levels of increasingly explicit prompts | Graded<br>-maximum 5 points per subtest<br>(3 subtests)<br>-higher score = worse | PA<br>-phoneme matching<br>Decoding<br>-word reading fluency | CTOPP<br>-sound matching | WR<br>NWR<br>PR | WRMT<br>-word attack<br>-word identification<br>-passage comprehensio n<br>TOWRE<br>-sight word efficiency<br>-phonemic decoding efficiency<br>CRAB<br>-oral reading fluency<br>Running Records |
| **Coventry, W.L., Byrne, B., Olson, R.K., Corley, R. & Samuelsson, S. (2011)** | PA<br>-identification of 4 phonemes in real words | Explicit Verbal | Graded<br>-4 levels of increasingly explicit prompts | -% correct score across the 4 subtests (6 items per subtest) | PA<br>-syllable & phoneme blending | Researcher developed tools | WR<br>NWR | TOWRE<br>-sight word efficiency |

| | | | | | | | | |
|---|---|---|---|---|---|---|---|---|
| | | | | | -phoneme matching<br>-phoneme deletion<br>-rhyme and final phoneme recognition<br><br>SSK<br>-Letter name knowledge<br>-Letter sound knowledge | | | -phonemic decoding efficiency |
| **Cunningham, A. & Carroll, J. (2011)** | PA<br>- phoneme segmentation nonwords | Not specified | Graded<br>- 7 levels of increasingly explicit prompts | Graded<br>-maximum 6 points per item<br>-high score = better | PA<br>- phoneme segmentation<br>- phoneme deletion<br><br>SSK<br>- letter-sound knowledge<br>-letter-name knowledge | PAT<br>-phoneme deletion<br><br>Researcher developed tools | WR<br>WW | BAS<br>-word reading<br>-word spelling/writi ng |

| | | | | | | | | |
|---|---|---|---|---|---|---|---|---|
| **Edwards, A. (2020)** | Decoding<br>-read nonwords<br>(CVC, CVCe) | Explicit Verbal | Graded<br>-5 levels of increasingly explicit prompts | Graded<br>-maximum 5 points per level<br>(2 levels)<br>-higher score = worse | PA<br>-phoneme deletion<br>-phoneme matching<br><br>Decoding<br>-timed word reading<br>-timed nonword reading<br>-untimed word reading<br>-untimed nonword reading | CTOPP<br>-elision<br>-sound blending<br><br>WRMT<br>-word identification<br>-word attack<br><br>TOWRE<br>-sight word efficiency<br>-phonemic decoding efficiency | PR | CRAB<br>-passage reading fluency |

| | | | | | | | | |
|---|---|---|---|---|---|---|---|---|
| **Gellert, A.S. & Elbro, C. (2017a)** | SSK<br>-learn novel symbol-sound corresponden ce<br><br>Decoding<br>-read nonwords comprised of 2 novel symbols<br>-read nonwords comprised of 3-4 novel symbols | Explicit verbal | N/A | -#of symbol-sounds produced correctly<br>-# of nonwords read correctly | PA<br>-phoneme identification<br>-phoneme blending<br><br>SSK<br>-letter name knowledge<br><br>Decoding<br>-word reading | Researcher developed tools | WR<br>NWR | Researcher developed tools |
| **Gellert, A.S. & Elbro, C. (2017b)** | PA<br>- phoneme identification real words | Explicit verbal | Graded<br>-3 levels of increasingly explicit prompts | Graded<br>-maximum 4 points per item<br>-higher score = better | PA<br>- phoneme identification<br><br>SSK<br>- letter name knowledge<br><br>Decoding | Researcher developed tools | WR<br>NWR | Researcher developed tools<br><br>Dynamic decoding measure (described above) |

| | | | | | | | | |
|---|---|---|---|---|---|---|---|---|
| | | | | | - word reading | | | |
| **Gillam, S.L., Fargo, J. Foley, B. & Olszewski, A. (2011)** | PA<br>-nonverbal phoneme deletion real words | Explicit verbal | Graded<br>-4 levels of increasingly explicit prompts | Graded<br>-maximum 5 points per item<br>-higher score = worse | PA<br>-phoneme deletion<br>Decoding<br>-read nonwords | Researcher developed tools<br>WRMT-R/NU<br>-word attack | N/A | N/A |
| **Hautala, J., Heikkila, R., Nieminen, L., Rantanen, V. Latvala, J-M. & Richardson,U. (2020)** | Decoding<br>-word reading accuracy and fluency<br>-nonword reading accuracy and fluency | Explicit visual and auditory feedback in game | N/A | -# of correct responses (accuracy)<br>-# of correct responses per minute (fluency) | Decoding<br>-word reading fluency<br>-nonword reading | Lukilasse 2<br>-word reading fluency<br>LukiMat<br>-pseudoword reading | N/A | N/A |
| **Horbach, J., Shcarke, W., Cröll, J., Heim, S. & Günther, T. (2015)** | SSK<br>-learn novel symbol-syllable | Explicit verbal (train phase) | N/A | -% of correct responses | PA<br>-rhyming | BISC<br>-rhyming words | WR<br>NWR | SLRT-II |

| | | | | | | | | |
|---|---|---|---|---|---|---|---|---|
| | correspondence<br><br>Decoding<br>-read nonwords comprised of novel symbols | No feedback (test phase) | | | -syllable segmentation<br>-phoneme identification<br>-phoneme blending<br><br>Decoding<br>-read nonsense syllables | -counting syllables<br>-first sound identification<br>-blending phonemes<br><br>Researcher developed tools | | -word reading accuracy and speed<br>-nonword reading accuracy and speed |
| **Horbach, J. Weber, K. Opolony, F. Scharke, W. Radach, R. Heim, S. & Günther, T. (2018)** | SSK<br>-learn novel symbol-syllable correspondence | Explicit verbal (train phase)<br><br>No feedback (test phase) | N/A | -% of correct responses | SSK<br>-letter name or sound knowledge | Researcher developed tools | WR<br>NWR | SLRT-II<br>-word reading speed and accuracy<br>-nonword reading speed and accuracy |
| **Law, J.M., De Vos, A., Vanderauwera, J., Wouters, J. Ghesquiere, P/ & Vandermosten, M. (2018)** | SSK<br>-learn novel symbol-sound correspondence<br><br>Decoding<br>-read real words | Implicit<br>-correct responses lead to game success | N/A | SSK<br>-number of correct responses<br>-speed of response<br><br>Decoding<br>–number of words read | PA<br>-phoneme deletion<br><br>SSK<br>-orthographic knowledge | Researcher developed tools<br>-phoneme deletion<br>-orthographic knowledge choice task | N/A | N/A |

| | | | | | | | | |
|---|---|---|---|---|---|---|---|---|
| | comprised of novel symbols | | | correctly per second | judgement task<br><br>Decoding<br><br>-word reading speed<br><br>-nonword reading speed | EMT<br><br>-word reading fluency<br><br>Klepel<br><br>-nonword reading fluency | | |
| **Liu, C., Hoa Chung, K.K., Wang, L.C. & Liu, D. (2021)** | SSK<br><br>-learn novel symbol-sound corresponden ce | Explicit verbal<br><br>(train phase)<br><br>No feedback<br><br>(test phase) | N/A | -# of correct responses | PA<br><br>-syllable deletion<br><br>SSK<br><br>-Chinese orthographic knowledge judgement task<br><br>Decoding<br><br>-one- and two-character Chinese word reading | Researcher developed tools | N/A | N/A |

| | | | | | | | | |
|---|---|---|---|---|---|---|---|---|
| **Lu, Y. & Hu, C (2019)** | PA<br>- phoneme segmentation nonwords | Explicit verbal and visual (pennies in Elkonin boxes) | Graded<br>- 7 levels of increasingly explicit prompts | Graded<br>- maximum 6 points per item<br>-higher score =better | PA<br>- phoneme segmentation | Researcher developed tools | N/A | N/A |
| **Osa Fuentes, P.M. (2003)** | PA<br>-syllable isolation real words<br>-syllable matching real words<br>-invented spelling real words | Explicit verbal | Graded<br>-4 levels of increasingly explicit prompts | Graded<br>-maximum 5 points per item | PA<br>-syllable isolation real words<br>-syllable matching real words<br>-invented spelling real words | Yopp Singer modified tasks<br>Mann & Colb (1987) modified task | WR | Kaufman test of decoding (K-ABC) |
| **Petersen, D. B., & Gillam, R. B. (2015)** | Decoding<br>-read nonwords (CVC) | Explicit verbal (train phase) | N/A | Graded<br>-decoding strategy score<br>-maximum 4 points<br>-higher score = better<br>Graded | PA<br>-phoneme identification<br>SSK<br>-letter name knowledge | DIBELS<br>-initial sound fluency<br>-letter naming fluency | WR<br>NWR<br>PR | WRMT-R<br>-word identification<br>DIBELS<br>-nonword fluency<br>-oral reading fluency |

| | | | | | | | | |
|---|---|---|---|---|---|---|---|---|
| | | | | -response to intervention score<br>-maximum 2 points<br>-higher score = better<br><br>Total Modifiability score<br>-decoding strategy + response to intervention score<br>-maximum 6 points<br>-higher score = better | | | | |
| **Petersen, D.B, Allen, M.M. & Spencer, T.D. (2016)** | Decoding<br>-read nonwords<br>(CVC) | Explicit verbal (train phase) | N/A | Graded<br>-learning rating scale<br>-maximum 4 points<br>-higher score = better | PA<br>-phoneme identification<br><br>SSK<br>-letter name knowledge | DIBELS<br>-initial sound fluency<br>-letter naming fluency | WR<br>NWR | TOWRE<br>-sight word reading efficiency<br>-phonemic decoding efficiency |

| | | | | | | | | |
|---|---|---|---|---|---|---|---|---|
| | | | | Graded<br>-post-test strategy score<br>-maximum 3 points per item<br>(4 items)<br>-higher score = better | | | | DIBELS<br>-nonsense word fluency |
| | | | | Total composite score<br>-learning rating + post-test strategy scores<br>-maximum 16 points<br>-higher score = better | | | | |
| **Sittner Bridges, M. & Catts, H.W. (2011a)**<br>**(Sample 1)** | PA<br>-syllable & phoneme deletion real words | Explicit verbal | Graded<br>-3 levels | Graded<br>-maximum 4 points per item -higher sore = better | PA<br>-syllable & phoneme deletion | SDT<br>-syllable & phoneme deletion | WR<br>NWR | WRMT-R/NU<br>-word identification<br>-word Attack |

| | | | | | | | | |
|---|---|---|---|---|---|---|---|---|
| **Sittner Bridges, M. & Catts, H.W. (2011b)**<br><br>**(Sample 2)** | PA<br><br>-syllable and phoneme deletion real words | Explicit verbal | Graded<br><br>- 3 levels | Graded<br><br>-maximum 4 points per item<br><br>-higher score = better | PA<br><br>-phoneme identification | DIBELS<br><br>-initial sound fluency | WR<br><br>NWR | WRMT-R/NU<br>-word identification<br><br>-word Attack |
| **Spector, J. (1992)** | PA<br><br>-phoneme segmentation real words | Explicit verbal | Graded<br><br>-7 levels | Graded<br><br>-maximum 6 points per item<br><br>-higher score = better | PA<br><br>-phoneme segmentation<br><br>-phoneme deletion | Yopp Singer Task<br><br>-phoneme segmentation<br><br>Bruce Word Analysis Test<br><br>-phoneme deletion | WR<br><br>WW | San Diego Quick Assessment List<br><br>-word recognition<br><br>Invented Spelling<br><br>-word writing |
| **Teeuwen, E. (2020)** | SSK<br><br>- learn novel symbol-sound correspondence | Implicit visual<br><br>-correct responses lead to game success | N/A | Accuracy score<br><br>-# of correct responses<br><br>Speed score<br><br>-average response time | PA<br><br>-phoneme identification<br><br>SSK<br><br>-letter name knowledge | Researcher developed tools<br><br>-first sound awareness task<br><br>-letter knowledge task | N/A | N/A |

| | | | | | | | | |
|---|---|---|---|---|---|---|---|---|
| **Yap, D.F-F. (2018)** | PA<br>-phoneme segmentation real words | Explicit verbal | Graded<br>-7 levels | Graded<br>-maximum 6 points per item<br>-higher score = better | PA<br>-phoneme deletion<br>-phoneme blending<br>-phoneme identification<br>SSK<br>-letter sound knowledge<br>Decoding<br>-word reading | CTOPP-2<br>-elision<br>-phoneme blending<br>-sound matching<br>YARC<br>-letter sound knowledge<br>-early word recognition | WR<br>NWR | WJ-III<br>-letter-word identification<br>-word attack |

| | | | | | | | | |
|---|---|---|---|---|---|---|---|---|
| **Zumeta, R.O. (2010)** | PA<br>-phoneme segmentation real words | Explicit verbal | | Graded<br>-maximum 6 points 6 across 5 levels of instruction<br>-higher score = better | PA<br>-phoneme deletion<br>-phoneme blending<br>-phoneme identification<br><br>Decoding<br>-word reading | DIBELS<br>-phoneme segmentation fluency<br>-phoneme segmentation accuracy<br><br>CTOPP<br>-elision<br>-phoneme blending<br>-sound matching<br><br>Researched developed tools<br>-word identification fluency | N/A | N/A |

*Note.* DA=dynamic assessment, SA= static assessment, PA = phonological awareness, SSK = sound-symbol knowledge, WR = word reading, NWR= non-word reading, BAS=Benchmark Assessment System, BISC=Bielefelder screening, CRAB= Comprehensive reading assessment battery, CTOPP= Comprehensive test of phonological processing, CV= Consonant Vowel, DIBELS = Dynamic indicators of basic early literacy skills, FSF = First sound fluency, LNF= Letter naming fluency, PAT=Phonological awareness test, SLRT-III=Salzburger lese und rechtschreibtest, SWRT=Sight word reading test, TOWRE= Test of word reading efficiency, WJ-III = Woodcock Johnson-III, WIAT=Wechsler individual achievement test, WRAT=, WRMT(R/NU)=Woodcock reading mastery tests, YARC= York assessment of reading comprehension

**Table 3.**

***Quality Appraisal of Included Studies***

| Study | Participants | Dynamic Assessment | | Static Assessment &/ WR Outcome Measures | | Flow and Timing | | Statistical Analyses | Overall Score and /12 & % | Overall Appraisal |
|---|---|---|---|---|---|---|---|---|---|---|
| | *Were participant characteristics adequately described?* (1) | *Was the DA adequately described?* *(2)* | *Was the test appropriate for evaluating early literacy skills?* *(2)* | *Was the SA or WR outcome measure adequately described?* *(2)* | *Was the SA or OM valid and reliable for evaluating early literacy skills?* (2) | *Were all participants included in all analyses?* (1) | *If not, were reasons for exclusion or loss to follow up adequately described?*(1) | *Were appropriate statistical tests used?* (1) | | |
| **Aravena, S., Snellings, P., Tijms, J., & van der Molen, M.W. (2013)** | 1 | 2 | 2 | 2 | 2 | 0 | 1 | 1 | 11(92%) | High |
| **Aravena, S., Tijms, J. Snellings, P. & van der Molen, M.W. (2018)** | 1 | 2 | 1 | 2 | 2 | 1 | 1 | 1 | 11(92%) | High |
| **Barker, R.M. & Saunders, K.J. (2020)** | 1 | 1 | 2 | 2 | 2 | 0 | 1 | 1 | 10(83%) | High |
| **Caffrey, E. (2006)** | 1 | 2 | 2 | 2 | 2 | 0 | 1 | 1 | 11(92%) | High |
| **Catts, H.W., Nielsen, D.C., Sittner Bridges, M., Liu, Y. S., &** | 1 | 2 | 2 | 2 | 2 | 0 | 1 | 1 | 11(92%) | High |

| | | | | | | | | | | |
|---|---|---|---|---|---|---|---|---|---|---|
| **Bontempo, D.E. (2015)** | | | | | | | | | | |
| **Cho, E. & Compton, D.L. (2015)** | 1 | 2 | 2 | 2 | 2 | 1 | 1 | 1 | 12(100%) | High |
| **Cho, E., Compton, D., Fuchs, D., Fuchs, L.S. & Bouton, B. (2014)** | 1 | 2 | 2 | 2 | 2 | 0 | 1 | 1 | 11(92%) | High |
| **Cho, E., Compton, D.L., Gilbert, J.K., Steacy, L.M., Collins, A.A. & Lindstrom, E.R. (2017)** | 1 | 2 | 2 | 2 | 2 | 0 | 1 | 1 | 11(92%) | High |
| **Compton, D. L., Fuchs, D., Fuchs, L. S., Bouton, B., Gilbert, J. K., Barquero, L. A., Cho, E., Crouch, R.C. (2010)** | 1 | 2 | 2 | 2 | 2 | 0 | 1 | 1 | 11(92%) | High |
| **Coventry, W.L., Byrne, B., Olson, R.K., Corley, R. & Samuelsson, S. (2011)** | 1 | 2 | 2 | 2 | 2 | 0 | 0 | 1 | 10(83%) | High |

| | | | | | | | | | | |
|---|---|---|---|---|---|---|---|---|---|---|
| **Cunningham, A. & Carroll, J. (2011)** | 0 | 2 | 2 | 2 | 2 | 1 | 1 | 1 | 11(92%) | High |
| **Edwards, A. (2020)** | 0 | 0 | 2 | 2 | 2 | 1 | 1 | 1 | 9(75%) | High |
| **Gellert, A.S. & Elbro, C. (2017a)** | 1 | 2 | 2 | 2 | 2 | 0 | 1 | 1 | 11(92%) | High |
| **Gellert, A.S. & Elbro, C. (2017b)** | 1 | 2 | 2 | 2 | 2 | 0 | 1 | 1 | 11(92) % | High |
| **Gillam, S.L., Fargo, J. Foley, B. & Olszewski A. (2011)** | 1 | 2 | 2 | 2 | 2 | 1 | 1 | 1 | 12(100%) | High |
| **Hautala, J., Heikkila, R., Nieminen, L., Rantanen, V. Latvala, J-M. & Richardson,U. (2020)** | 1 | 2 | 2 | 2 | 2 | 0 | 1 | 1 | 11(92%) | High |
| **Horbach, J. Weber, K. Opolony, F. Scharke, W. Radach, R. Heim, S. & Günther, T. (2018)** | 1 | 2 | 2 | 2 | 2 | 0 | 1 | 0 | 10(83%) | High |

| | | | | | | | | | | |
|---|---|---|---|---|---|---|---|---|---|---|
| **Horbach, J., Shcarke, W., Cröll, J., Heim, S. & Günther, T. (2015)** | 1 | 2 | 2 | 2 | 2 | 0 | 0 | 1 | 10(83%) | High |
| **Law, J.M., De Vos, A., Vanderauwera, J., Wouters, J. Ghesquiere, P/ & Vandermosten, M. (2018)** | 1 | 2 | 2 | 2 | 2 | 1 | 1 | 1 | 12(100%) | High |
| **Liu, C., Hoa Chung, L.C., Wang, L.C. & Liu, D. (2021)** | 1 | 1 | 2 | 2 | 0 | 1 | 1 | 1 | 9(75%) | High |
| **Lu, Y. & Hu, C (2019)** | 0 | 2 | 2 | 2 | 2 | 1 | 1 | 1 | 11(92%) | High |
| **Osa Fuentes, P.M. (2003)** | 0 | 2 | 2 | 2 | 0 | 1 | 1 | 1 | 9(75%) | High |
| **Petersen, D. B., & Gillam, R. B. (2015)** | 1 | 2 | 2 | 2 | 2 | 1 | 1 | 1 | 12 (100%) | High |
| **Petersen, D.B, Allen, M.M. & Spencer, T.D. (2016)** | 0 | 2 | 2 | 2 | 2 | 1 | 1 | 1 | 11(92%) | High |
| **Sittner Bridges, M. & Catts, H.W. (2011)** | 0 | 2 | 2 | 2 | 2 | 1 | 1 | 1 | 11 (92%) | High |

| | | | | | | | | | | |
|---|---|---|---|---|---|---|---|---|---|---|
| **Spector, J. (1992)** | 1 | 2 | 2 | 2 | 2 | 0 | 1 | 1 | 11(92%) | High |
| **Teeuwen, E. (2010)** | 1 | 2 | 2 | 2 | 0 | 0 | 1 | 0 | 8(67%) | Medium |
| **Yap, D.F-F. (2018)** | 1 | 2 | 2 | 2 | 2 | 0 | 1 | 1 | 11(92%) | High |
| **Zumeta, R.O. (2010)** | 1 | 2 | 2 | 2 | 2 | 1 | 1 | 1 | 12(100%) | High |

*Note.* DA= Dynamic assessment, SA= Static assessment, WR = word reading, Low = 0-33%, Medium=34-66%, High=67-100%

### Appendix 3.

**Figure 10.**

*Baujat plot for studies included in the meta-analysis of the concurrent validity of dynamic and static assessments of early literacy skills*

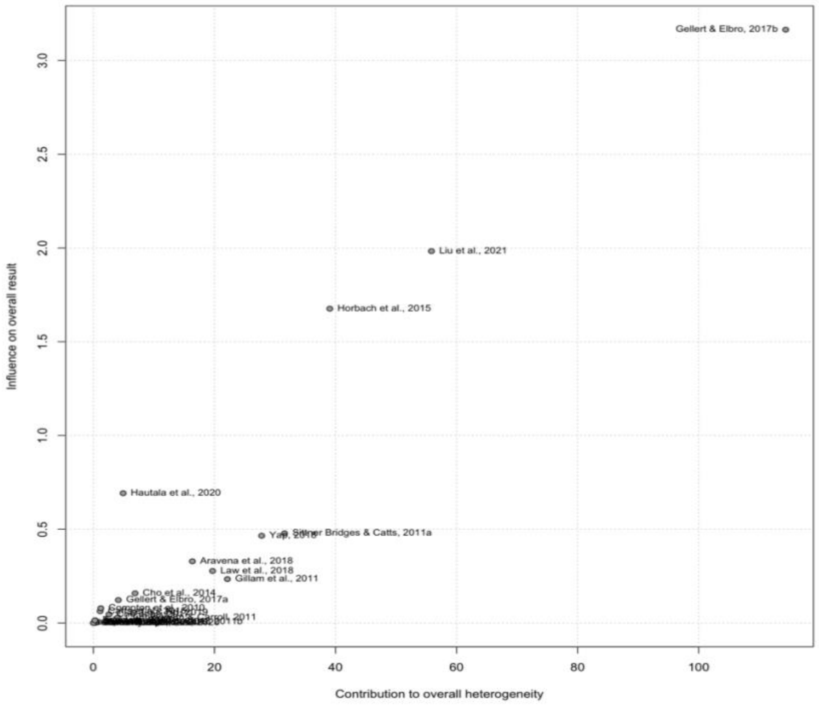


*Note.* In the Baujat plot, individual contribution to overall heterogeneity is represented on the horizontal axis, and the influence on overall result on the vertical axis. Studies with greates influence are found in the top right quadrant of the figure. Drawn in R using the ‘metacor’ package (R Core Team, 2021; Laliberté, 2019).

**Figure 11.**

*Baujat plot for studies included in the meta-analysis of the predictive validity of dynamic assessments of early literacy skills with word reading outcome measures*

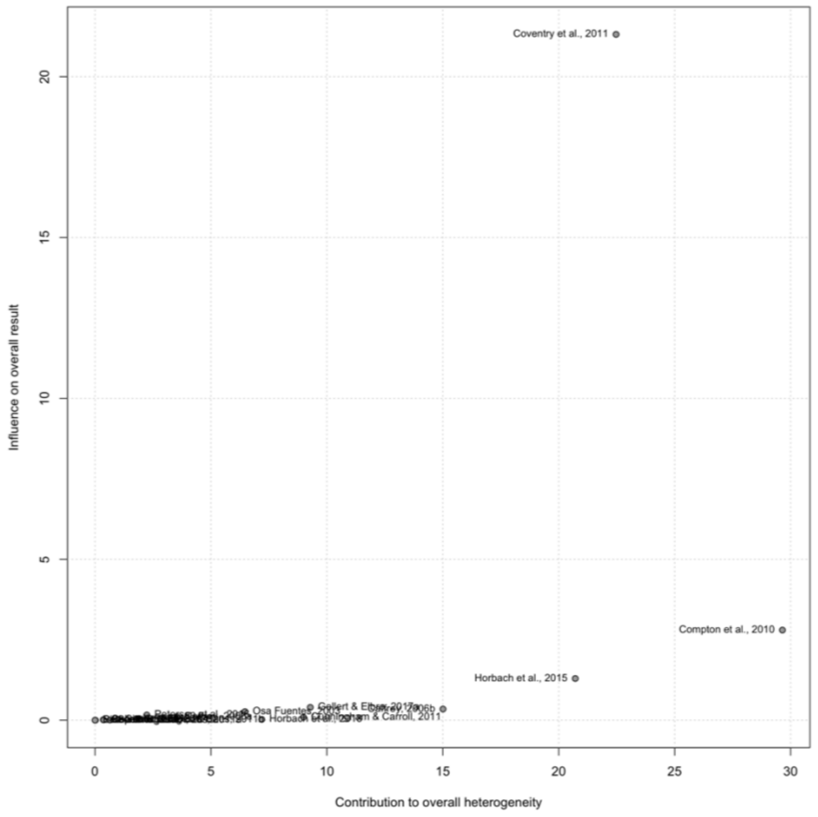


*Note*. In the Baujat plot, individual contribution to overall heterogeneity is represented on the horizontal axis, and the influence on overall result on the vertical axis. Studies with greatest influence are found in the top right quadrant of the figure. Drawn in R using the ‘metacor’ package (R Core Team, 2021; Laliberté, 2019).
